# Characterizing spatiotemporal white matter hyperintensity pathophysiology in vivo to disentangle vascular and neurodegenerative contributions

**DOI:** 10.1101/2025.06.10.25329342

**Authors:** Olivier Parent, Zaki Alasmar, Sophia Osborne, Aurélie Bussy, Manuela Costantino, Jérémie P. Fouquet, Daniela Quesada, Alexandre Pastor-Bernier, Alfonso Fajardo-Valdez, Alexa Pichet-Binette, Ann McQuarrie, Josefina Maranzano, Gabriel A. Devenyi, Christopher J. Steele, Sylvia Villeneuve, the PREVENT-AD Research Group, the Alzheimer’s Disease Neuroimaging Initiative (ADNI), Mahsa Dadar, M. Mallar Chakravarty

## Abstract

White matter hyperintensities (WMHs) are neuroimaging markers widely interpreted as caused by cerebral small vessel disease, yet emerging evidence suggests that a subset may have a neurodegenerative etiology. Current imaging methods have lacked the specificity to disentangle biological processes underlying WMHs *in vivo*. Here, we used voxel-level normative modeling and seven microstructural MRI markers with complementary biophysical sensitivities to generate single-subject high-resolution WMH pathophysiology maps in a large cohort (*n*=32,526). We calculated data-driven spatial patterns of similar WMHs, revealing distinct periventricular, posterior, and anterior clusters. We identified a reproducible WMH signature linked to dementia and Alzheimer’s disease, characterized by a posterior predominance and a pathophysiological pattern indicative of selective fiber degeneration. Posterior WMHs connected cortical regions vulnerable to tau pathology. Our framework distinguishes vascular and neurodegenerative contributions of WMHs *in vivo*, which could alter the course of treatment strategies and provide nuanced interpretations of research findings.

## Introduction

White matter hyperintensities (WMHs) of presumed vascular origins are among the most common brain radiological findings in older adults.^1,2^ They appear in most individuals over 65 years old and are typically detected as bright regions in fluid-attenuated inversion recovery (FLAIR) images during magnetic resonance imaging (MRI) exams.^3^ The presence of WMHs is widely considered a downstream consequence of cerebral small vessel disease.^4^ The extent of visible WMHs is used by neurologists and radiologists to monitor the cerebrovascular disease burden and for diagnoses such as vascular dementia,^5^ by researchers as a quantifiable marker of cerebrovascular dysfunction,^6,7^ and in some clinical trials as outcome measures of vascular treatments (e.g., antihypertensives).^8,9^ The nomenclature “of presumed vascular origins”^2^ highlights a key issue: the pathogenesis of WMHs can vary, with a growing body of evidence suggesting that some WMHs may be caused in part by neurodegenerative processes rather than solely vascular disease.^10–13^ For example, a post-mortem examination found no evidence of hypoperfusion-related damage within the WMHs of brain specimens with Alzheimer’s disease pathology, and instead observed signs of Wallerian degeneration.^11^ FLAIR images lack the specificity to parse the biological processes underlying WMHs, as these contain various levels of inflammation,^14^ edema,^15,16^ demyelination,^11,17^ and axonal degeneration^11^ that cannot be distinguished with a single contrast. A multimodal *in vivo* characterization of the underlying WMH pathophysiology is key in identifying WMHs that are specifically associated with neurodegenerative conditions. Having an improved understanding of WMH pathogenesis is crucial to avoid misinterpretations of cerebral pathology, which could influence both clinical management and research findings.

There is evidence that some of the pathophysiological heterogeneity observed in WMHs may be dependent on their neuroanatomical location. Studies suggest that periventricular WMHs are associated to a larger extent with vascular risk factor burden and contain more edema and less demyelination relative to deep WMHs.^18^ Consequently, periventricular and deep WMHs are often assessed separately using heuristic methods.^19^ This practice may introduce confirmation bias, since alternative parcellation schemes are rarely explored. To date, no study has used unbiased, data-driven methods to systematically investigate spatial patterns of WMH pathophysiology. Furthermore, the pathophysiological cascade of WMHs remains speculative, and it is unclear whether pathological events follow the same sequence in vascular and neurodegeneration-related WMHs. The prevailing hypothesis suggests that fluid infiltration from ventricles and blood vessels triggers inflammation, leading to tissue damage such as demyelination, oligodendrocyte loss, and, in more severe cases, axonal degeneration.^3^ Generally, WMH pathophysiology is characterized *post-mortem* using histological and immunohistochemical stains sensitive to different aspects of cellular, molecular, and pathological features. Multi-contrast microstructural MRIs provide markers with complementary biological sensitivities that can act as *in vivo* homologs of *post-mortem* stains,^20,21^ enabling the non-invasive, large-scale investigation of WMH pathophysiology and potentially providing clinicians with supplemental information about WMH pathogenesis that could alter the course of treatment and disease prevention strategies.

In this study, we generated high-resolution maps of WMH pathophysiology by performing voxel-wise normative modeling of seven MRI markers sensitive to fluid, fiber, myelin, and iron content in a large population cohort (UK Biobank; *n*=32,526) and transferred these models to an independent cohort of subjects along the Alzheimer’s disease spectrum. We calculated spatial regions of WMHs that showed similar pathophysiological features with unsupervised clustering, allowing us to then estimate region-specific pathophysiological cascades with machine learning-based disease modeling techniques. With an improved understanding of these spatiotemporal dynamics, we identified reproducible disease-specific signatures of WMH pathophysiology in vascular and neurodegenerative disorders.

## Results

### Image processing

We used data from 32,526 UK Biobank (UKB) participants, a population-based prospective study of middle- to late-aged individuals. Exclusion criteria (Methods) included disrupted MRI data used in downstream analyses (missing modality, motion artifacts or processing fails), missing demographic information, and a diagnosis of multiple sclerosis to not confound white matter lesions from WMHs (step-by-step exclusions detailed in Supplementary Fig. 1; demographic statistics detailed in Supplementary Table 1). While MRI images have already been processed by UKB,^22^ we updated key processing steps to ensure the highest data quality possible for our specific use case. A customized UKB template was generated^23^ (available at https://github.com/CoBrALab/WMH_patho_UKB) and MRI images were aligned to this template with multispectral registration (Methods), providing highly accurate voxel-by-voxel correspondence across subjects, particularly in the periventricular white matter which is an important area of interest. Second, to ensure accurate delineation of WMHs and normal-appearing white matter (NAWM) while leveraging multi-contrast information, we retrained the BraIn SegmentatiON (BISON) algorithm^24^ with a manually defined reference.

To investigate tissue properties, the multi-shell diffusion MRI and two-echo susceptibility-weighted MRI acquisitions were used to derive seven quantitative microstructural markers characterizing important aspects of white matter biophysical composition that are altered in aging, broadly categorized as fluid-sensitive (mean diffusivity [MD] and isotropic volume fraction [ISOVF]), fiber-sensitive (fractional anisotropy [FA], intracellular volume fraction [ICVF], and orientation dispersion [OD]), and myelin- and iron-sensitive (T2* and quantitative susceptibility mapping [QSM]). However, their biological underpinnings are complex and potentially disease-dependent (summarized in Table 1). By interpreting these markers together, we can better infer potential biological processes driving the observed effects (e.g., inflammation, axonal damage, edema, etc.).^21^

**Table 1.** Literature review of biological sensitivities of MRI microstructural markers. The main biological sensitivities listed here are a general guide only and should not be interpreted as the only sources of signal. In other words, the metrics are sensitive but not specific to the biological sources of signal listed here. The directionality of the associations is indicated.

| Modality | Marker | Physical interpretation | Biological sensitivities |
| --- | --- | --- | --- |
| Diffusion tensor imaging (DTI): Eigendecomposition of the diffusion signal resulting in a tensor described by three orthogonal axes of diffusion | Mean diffusivity (MD) | Average of diffusion along the three orthogonal diffusion axes of the tensor | Extracellular free water (+),<sup>25</sup> inflammation (+)<sup>26,27</sup> |
|  | Fractional anisotropy (FA) | Amplitude of diffusion in the principal direction relative to the other two axes | Intra-axonal water (+),<sup>25</sup> fiber organization (+)<sup>25</sup> |
| Neurite orientation dispersion and density imaging (NODDI): Modeling of the diffusion signal in three different tissue compartments | Isotropic volume fraction (ISOVF) | Fraction of the signal that diffuses completely isotropically | Extracellular free water (+)<sup>28,29</sup> |
|  | Intracellular volume fraction (ICVF) | Fraction of the anisotropic signal with negligible perpendicular diffusion (modeled as a collection of sticks) | Intra-axonal water (+)<sup>28,29</sup> |
|  | Orientation dispersion (OD) | Distribution of diffusion directions (0 when fibers are completely aligned, 1 when there is no preferred diffusion direction) | Fiber organization (-)<sup>28-30</sup> |
| Susceptibility-weighted imaging (SWI) | T2* | Exponential decay of the transverse relaxation impacted by susceptibility variations and spin-spin interactions | Myelin (-),<sup>31,32</sup> iron (-),<sup>31,32</sup> extracellular free water (+)<sup>33</sup> |
|  | Quantitative susceptibility mapping (QSM) | Sum of paramagnetic and diamagnetic susceptibilities | Myelin (-),<sup>31</sup> iron (+)<sup>31</sup> |

### Estimates of WMH pathophysiology

Pathophysiology was operationalized as the deviation in microstructural values in pathological tissue (WMHs) relative to the expected microstructure in healthy white matter. In other words, we aimed to estimate pathophysiological processes such as water accumulation/edema as opposed to microstructural properties like water density. High-resolution normative modeling of white matter microstructure was performed for the seven markers only including individual-level voxels that were labeled as NAWM (Methods; Fig. 1A),^34^ thus generating age- and sex-specific maps of expected healthy microstructure which are then used to normalize WMH microstructure using a z-score procedure (example in Fig. 1B). The resulting seven high-resolution pathophysiology maps represent in essence an individual-level signature of the biophysical processes at play within WMHs. These novel maps have many desirable properties: 1) they isolate the effect of the pathology within WMHs on the microstructure (i.e., the WMH pathophysiology), 2) since all markers are on the same scale, they do not require further normalization, and 3) they remove the spatial microstructural contrast driven by normal anatomical variations, which is a necessary property for our spatial clustering analyses (see below). As an example, the single-subject raw OD map in WMHs has a high level of spatial variation but these are not present in the z-scored map (Fig. 1B, first row compared to last row), highlighting that spatial variations were due to the anatomy and not caused by differences in WMH-related alterations.

**Figure 1.**
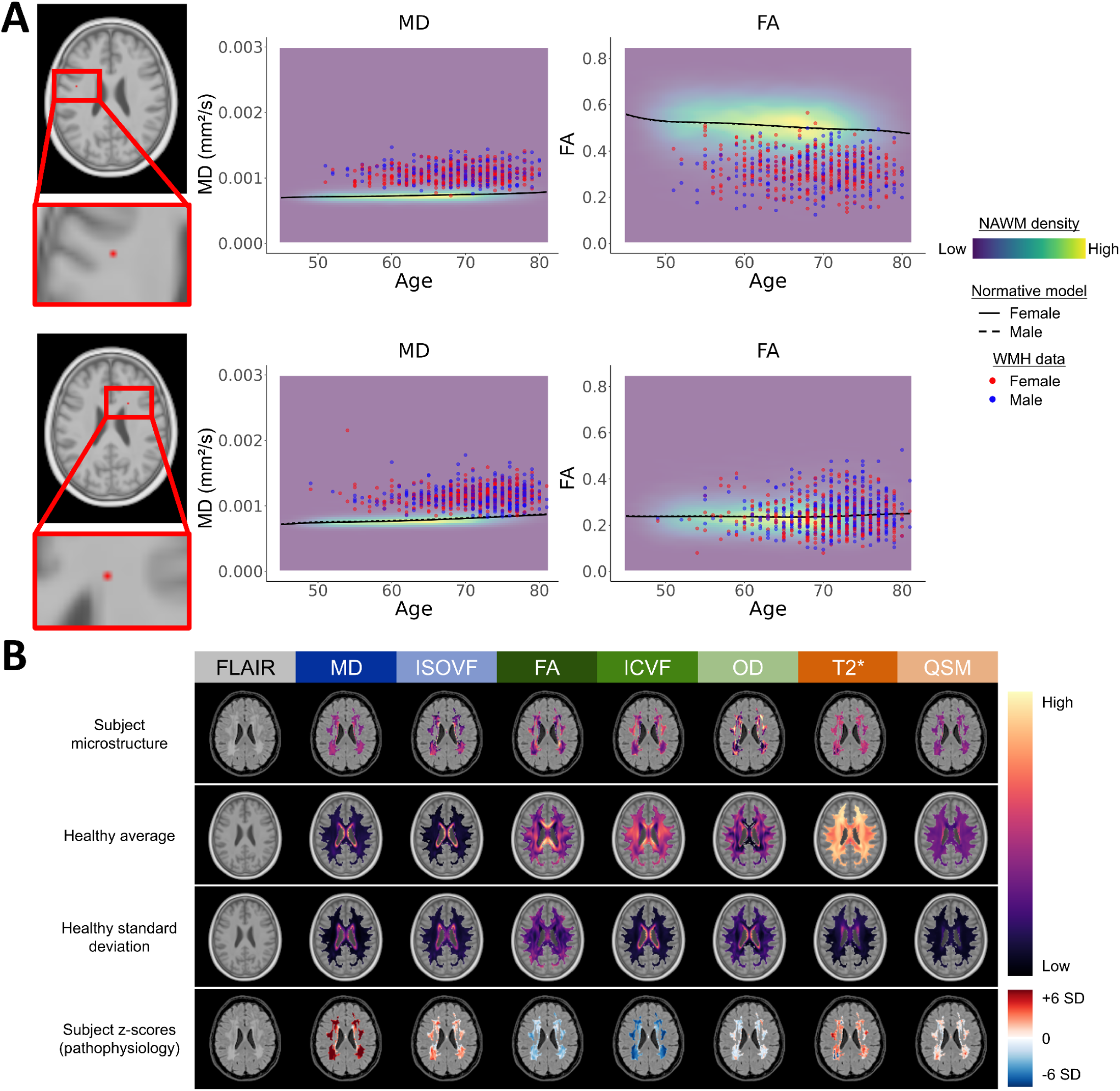
High-resolution estimates of WMH pathophysiology. **A)** Normative modeling of healthy microstructure was performed at each white matter voxel for the seven microstructural markers. The process is exemplified at two voxel locations for the MD and FA markers. The density of NAWM microstructural values, fitted normative models, and individual WMH microstructural data are shown. **B)** The procedure to derive WMH pathophysiological estimates is shown. First row: raw microstructural maps of one subject in UKB space. Second and third row: average and standard deviation maps of the expected healthy microstructure, matching the age and sex of the subject. Fourth row: abnormality z-scored maps (i.e., WMH pathophysiological estimates). Each microstructural marker is color-coded by its primary biological sensitivity: fluid-sensitive in shades of blue, fiber-sensitive in shades of green, and myelin and iron-sensitive in shades of orange. This color coding is kept throughout the manuscript.

### Spatial regions of pathophysiologically similar WMHs

From this rich dataset, we first aimed to characterize group-level spatial patterns of WMH pathophysiology in the general population. Subject-level maps of WMH pathophysiology were first collapsed into an average for each of the seven markers, excluding voxels with low WMH or NAWM prevalence (Fig. 2A), and were used as inputs to a spectral clustering algorithm.^35^ This data-driven approach identified spatial regions that are most similar between themselves solely in terms of WMH pathophysiology independent of voxel location. Yet, all clusters were mostly spatially contiguous, suggesting that WMHs appearing in specific areas shared pathophysiological signatures. We opted for a solution of three clusters (Fig. 2B) in the interest of parsimony since the four-cluster solution (Supplementary Fig. 2) was less spatially contiguous with similar pathophysiological differentiation across clusters. The first cluster surrounded the ventricular wall and showed low abnormality on all metrics (median z-scores: MD = 0.49, ISOVF = −0.18, FA = −0.61, ICVF = −1.26, OD = −0.3, T2* = 0.63, QSM = 0.06). The second cluster was located in periventricular-posterior regions and showed abnormality related to fluid accumulation, fiber alterations, and myelin and iron loss (see Table 1 for more complex biological interpretations; median z-scores: MD = 4.01, ISOVF = 1.07, FA = −1.56, ICVF = −3.22, OD = −0.3, T2* = 1.47, QSM = 0.23). The third cluster was located in deep-anterior regions and showed a similar pathophysiological pattern to the posterior cluster but with higher absolute abnormality on most metrics (median z-scores: MD = 5.89, ISOVF = 1.47, FA = −1.99, ICVF = −4.37, OD = −0.34, T2* = 1.6, QSM = 0.29).

**Figure 2.**
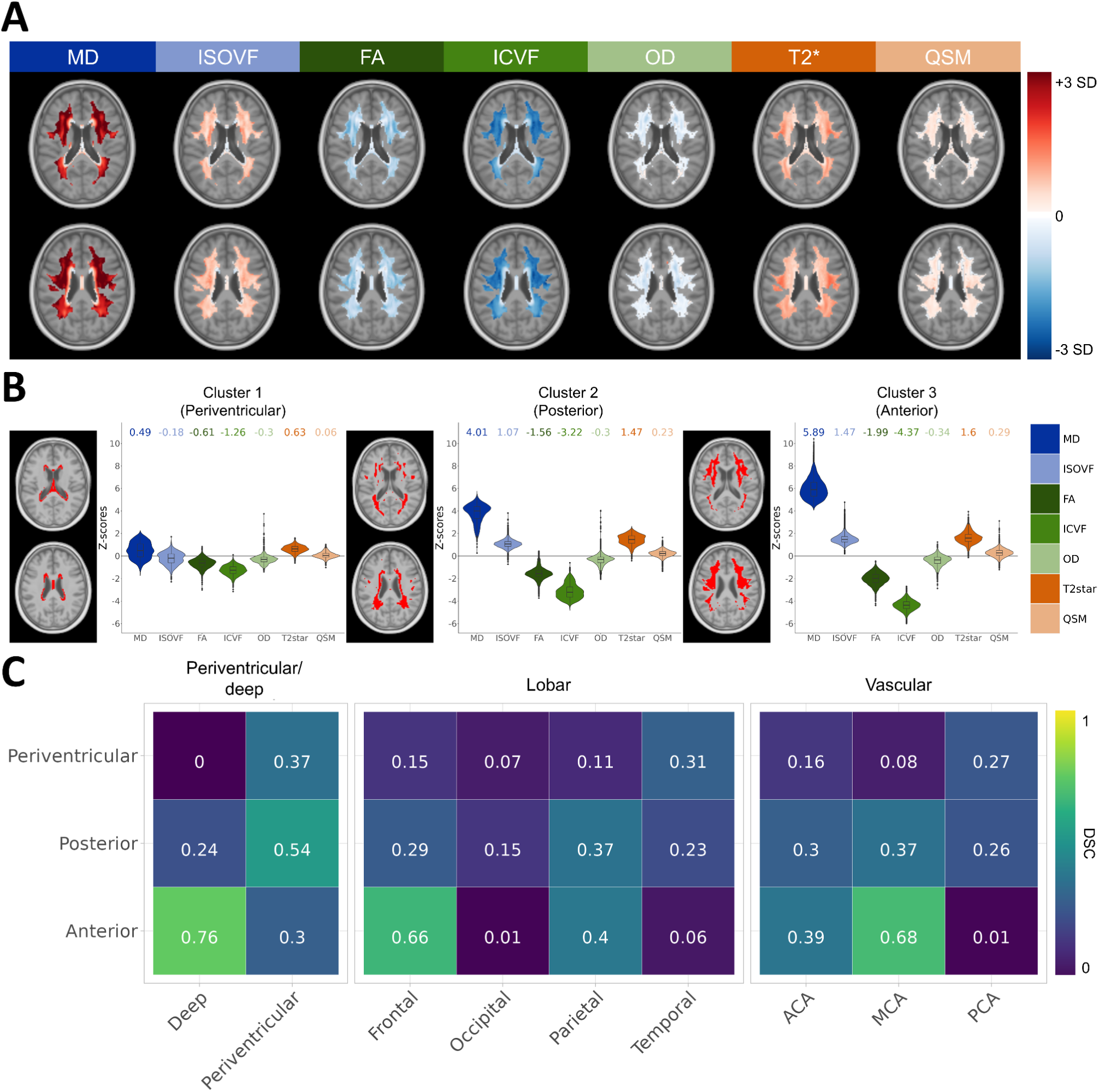
Spatial regions of pathophysiologically similar WMHs. **A)** Maps of between-subject averaged WMH pathophysiological estimates for each marker. Voxels with a low prevalence of WMH labels (<30) or NAWM labels (<5000) were excluded. These maps were used as inputs to a spectral clustering algorithm. **B)** Spatial clusters. Left: voxels included in the cluster are indicated in red. Right: Pathophysiological distributions of the voxels included in the cluster. Median values of those distributions are shown at the top. **C)** Dice similarity coefficients (DSCs) between our derived spatial clusters (y-axis) and WMH regions from other parcellations (x-axis). Only clustered voxels are included in the calculation. Abbreviations: anterior cerebral artery (ACA), middle cerebral artery (MCA), posterior cerebral artery (PCA)

To quantify and contextualize the localization of each spatial cluster, we compared the overlap of our novel parcellation with other heuristic parcellations used in the field: periventricular/deep, lobar regions, and cerebral artery territories as estimated from a probabilistic atlas (Fig. 2C).^36^ We observed that the first cluster was more periventricular (Dice similarity coefficient [DSC] = 0.37), located in temporal white matter (DSC = 0.31), and supplied by the posterior cerebral artery (DSC = 0.27). The second cluster was more periventricular (DSC = 0.54), located in parietal white matter (DSC = 0.37), and supplied by the middle cerebral artery (DSC = 0.37), although it also showed overlap with the posterior and anterior cerebral artery territories (DSC of 0.30 and 0.26, respectively). The third cluster was mostly located in deep and frontal white matter (DSC of 0.76 and 0.66, respectively) and largely supplied by the middle cerebral artery (DSC = 0.68). The lack of consistent agreement with often-used WMH parcellations suggests an improved pathophysiological specificity in these data-driven clusters that is not captured with the more heuristic approaches. For the rest of the manuscript, these spatial clusters will be referred to as periventricular (cluster 1), posterior (cluster 2), and anterior (cluster 3).

To investigate if these patterns simply reflected severity stages (e.g., spreading of WMHs from periventricular to posterior to anterior regions), we separated our sample into three groups based on total WMH volume and ran the clustering analysis within each group (Extended Fig. 1A). Remarkably, the spatial overlaps between the subgroup clusters and the original clustering solution in the complete dataset were very high (DSCs: Periventricular > 0.87; Posterior > 0.78; Anterior > 0.88; Extended Fig. 1B) and the associated pathophysiological patterns were highly congruent. We also found congruent spatial and pathophysiological patterns between males and females (DSCs: Periventricular > 0.95; Posterior > 0.91; Anterior > 0.96; Extended Fig. 2A-B). This demonstrated that the observed spatial variations in WMH pathophysiology are robust to the severity of WMH burden and are consistent across sexes.

Finally, voxels that were previously excluded from the clustering were assigned to the closest spatial cluster using a search area strategy (Methods; Supplementary Fig. 3). This novel parcellation enabled a pathophysiology-informed dimensionality reduction of the high-resolution WMH maps, as we extracted subject-wise region-of-interest (ROI) measures by sampling the median pathophysiological estimates within WMHs in each spatial cluster (Methods). To simplify the interpretation, we inverted the values of markers that showed negative effects in WMHs (FA, ICVF, and OD) so that an increase in z-scores is consistently interpreted as worsening WMH pathophysiology for all markers.

### Defining temporal pathophysiological cascades

Next, we investigated the temporal heterogeneity of WMH pathophysiology by modeling pathophysiological cascades in our three derived spatial regions using the Subtype and Stage Inference (SuStaIn) technique.^37^ SuStaIn is an event-based unsupervised machine learning technique that infers temporal progression patterns from cross-sectional data and is further able to find progression subtypes, essentially sorting abnormality events in the order in which they tend to appear across different clusters of subjects.^37^ Pathophysiological events were determined empirically on a per-marker basis (Methods; Supplementary Fig. 4).

Interestingly, we did not find evidence for sub-trajectories of WMH pathophysiological progression in any of the regions, as is evidenced by nearly identical biomarker progression curves even when investigating cross-validated three-trajectory solutions (Extended Fig. 3). We interpret this finding as evidence that most of the pathophysiological heterogeneity is spatial and not temporal, and was adequately accounted for in our spatial clustering step. We thus describe here the pathophysiological cascades shared across all subjects within each WMH region (Fig. 3A). Periventricular WMHs showed low pathophysiology across temporal stages, with ICVF being the earliest marker to be altered. Posterior WMHs displayed early alterations of the ICVF and MD markers, followed by FA, T2*, and ISOVF in the intermediate stages, and OD and QSM in the later stages. Anterior WMHs showed a similar progression pattern but with most markers reaching higher abnormality. Positional variance diagrams showed very low stage uncertainty, demonstrating the stability of these temporal trajectories (Supplementary Fig. 5). Using WMH volume as a simpler empirical definition of temporal disease severity, we recapitulate results from our data-driven findings of disease progression (Extended Fig. 4). The subject-level stage from SuStaIn provided a summary measure of the temporal progression of WMH pathophysiology and is included in further analyses.

**Figure 3.**
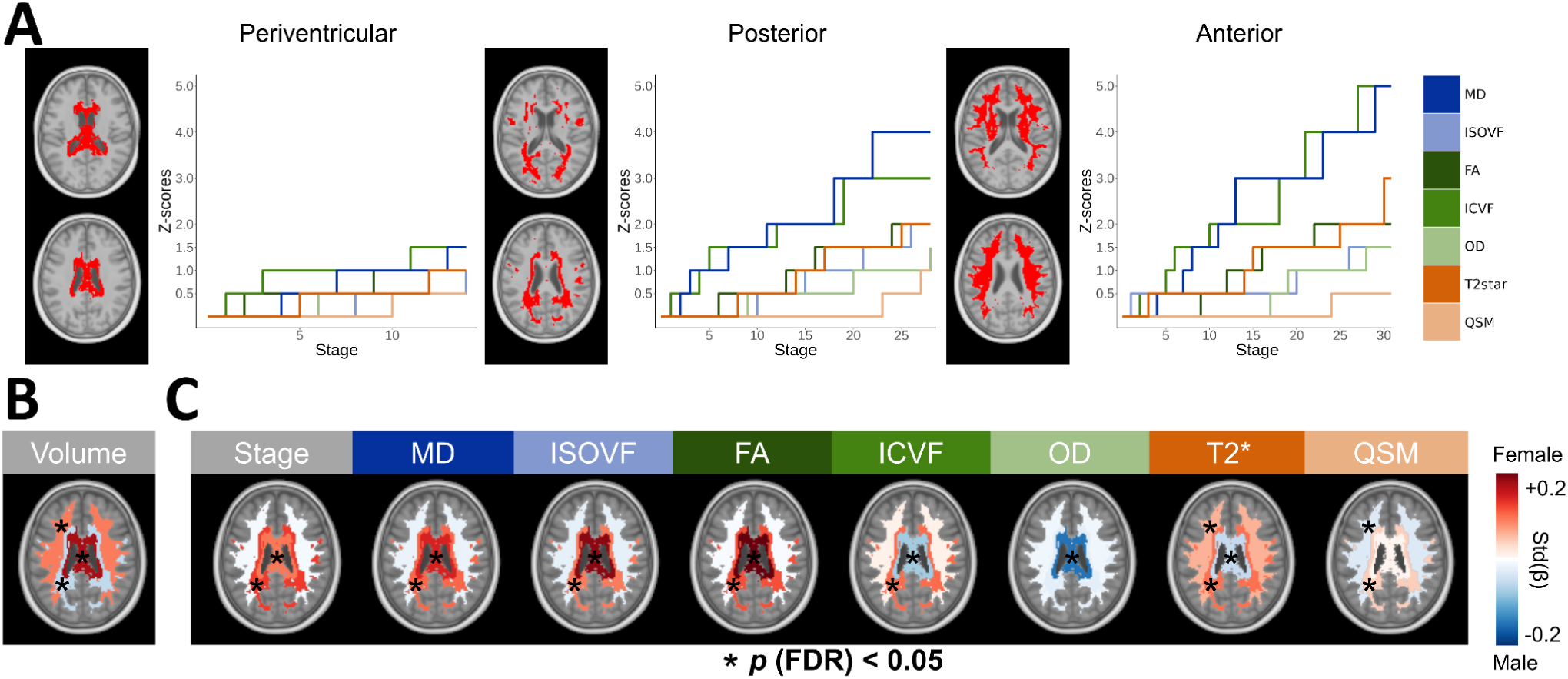
Pathophysiological cascades and sex differences. **A)** In each WMH spatial region, we modeled the temporal pathophysiological cascade using SuStaIn. Shown are the winner-take-all trajectories, with the x-axis representing a data-driven temporal axis of pathophysiological events (stages) and the y-axis representing the abnormality z-score thresholds. **B)** Sex differences in WMH volume. **C)** Sex differences in WMH pathophysiology, controlling for the corresponding regional WMH volume. Differences are expressed as standardized betas. Red colors indicate a higher effect in females and blue colors indicate a higher effect in males. Significant effects after performing false discovery rate (FDR) correction (*p*<0.05) are shown with asterisks.

### Sex differences in WMH pathophysiology

Previous studies have reported higher WMH burdens in females that manifest after midlife, which is hypothesized to be due to the menopausal transition and the consequent reduction in the neuroprotective effects of estrogen.^38,39^ We investigated whether a specific signature of WMH pathophysiology exists in females, providing standardized betas and false discovery rate (FDR)-corrected *p*-values.^40^ Interestingly, we found higher WMH volumes in the periventricular (*β*=0.17, *p* (FDR) < 0.001) and anterior (*β*=0.09, *p* (FDR) < 0.001) regions in females, but higher WMH volumes in posterior (*β*=-0.05, *p* (FDR) < 0.001) regions in males (Fig. 3B). Investigating differences in WMH pathophysiology when males and females have equivalent WMH volumes (Fig. 3C), we observed a clear pattern of strong effects in females restricted to periventricular and posterior WMHs for most pathophysiological markers. This was not the case for the OD marker which showed a significantly higher effect in males in the periventricular region but no significant effects in other regions. Taken together, our results show nuanced sex-specific effects in WMHs. There are clear spatial differences, with females having more WMHs but similar pathophysiological effects in anterior regions, while males show higher WMH volumes but lower pathophysiological effects in posterior regions.

### Pathophysiology of WMHs across vascular and neurodegenerative disorders

With a deeper understanding of WMH spatiotemporal dynamics, we were now well-positioned to elucidate complex disease-specific phenotypes in vascular and neurodegenerative disorders. We first compared WMHs across different diagnostic categories with a case-control design using UKB lifetime diagnosis data (i.e., diagnoses before or after the MRI visit; Fig. 4A). Control subjects had no endocrine, circulatory, behavioral, nervous, or metabolic disease-related lifetime diagnoses, with some exceptions (*n*=10,629; Methods). Case subjects were defined by groupings of ICD-10 codes (Supplementary Fig. 6).

**Figure 4.**
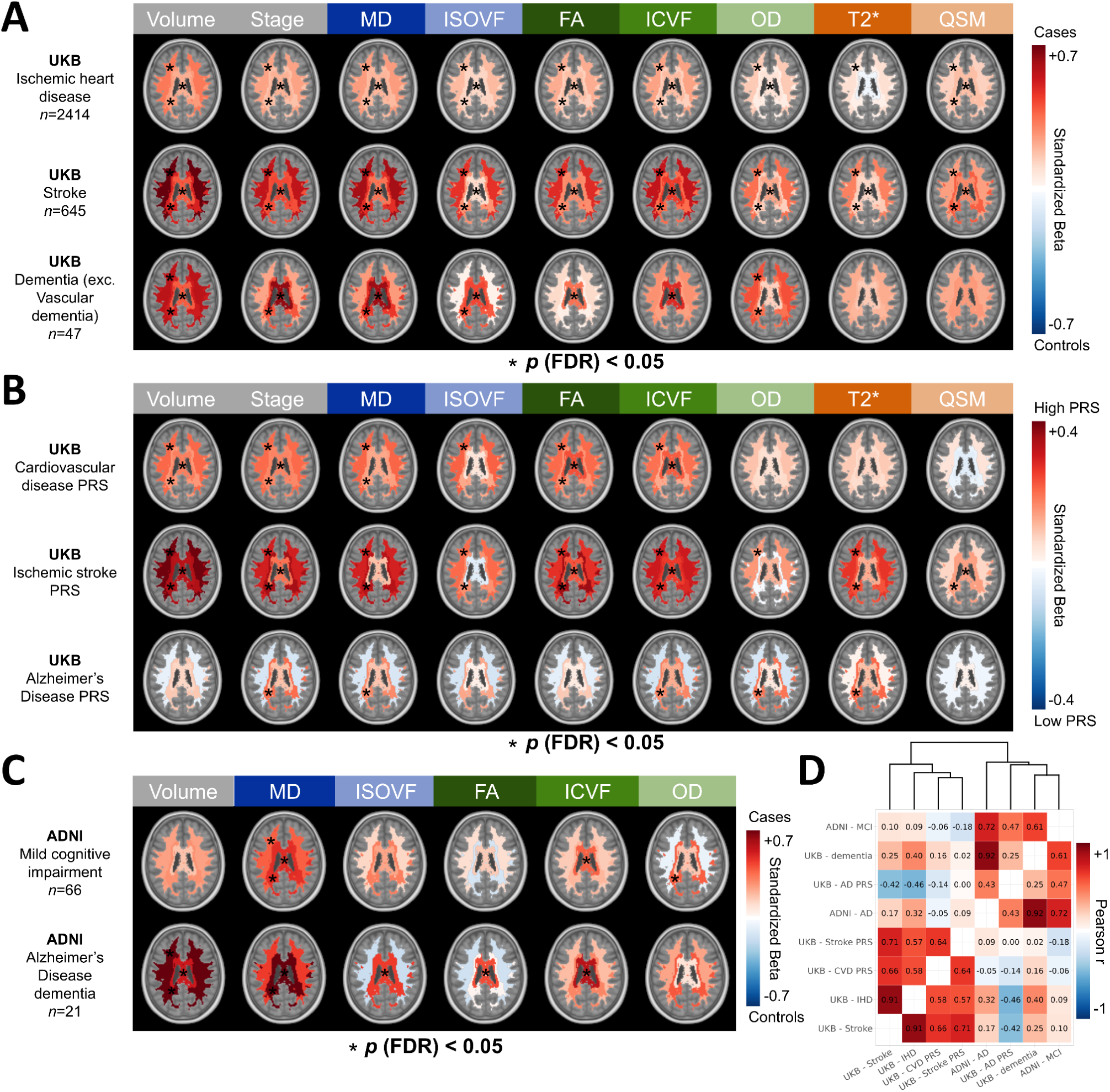
Differences in WMH pathophysiology across neurodegenerative and vascular diseases. Effect size patterns of spatiotemporal WMHs pathophysiology comparing cases and controls in **A)** UK Biobank participants according to ICD-10 code groupings of ischemic heart disease, stroke, and dementia, **B)** UK Biobank participants at high genetic risk of cardiovascular disease, ischemic stroke, and Alzheimer’s disease, and **C)** ADNI participants with mild cognitive impairment due to Alzheimer’s disease and Alzheimer’s disease dementia. Red colors indicate higher WMH burden in cases and blue colors indicate higher WMH burden in controls. Significant effects at the FDR-corrected *p*<0.05 level are indicated with black asterisks. **D)** Effect size patterns were correlated and clustered with hierarchical clustering.

In individuals with ischemic heart disease (*n*=2,414), WMH alterations were small but significant, with stronger effects in anterior regions (average *β* = 0.19; *p* (FDR) < 0.05 for all markers) than posterior regions (average *β* = 0.13; *p* (FDR) < 0.05 for all markers except OD and T2*). The stroke group (*n*=645) showed moderate-to-large effects, particularly in anterior WMHs (average *β* = 0.47; *p* (FDR) < 0.05 for all markers) relative to posterior WMHs (average *β* = 0.33; *p* (FDR) < 0.05 for all markers). In contrast, the dementia group excluding vascular dementia (*n*=47) showed moderate effects, particularly in posterior WMHs (average *β* = 0.31; *p* (FDR) < 0.05 for WMH volume, stage, MD, ISOVF, OD) relative to anterior WMHs (average *β* = 0.26; *p* (FDR) < 0.05 for WMH volume, OD). Notably, FA effects were more pronounced in stroke, whereas OD effects were more pronounced in dementia. This suggests a signature of neurodegenerative WMHs caused by specific disruptions to the fiber organization captured by the OD marker. However, this analysis is limited by the uncertainty in dementia type (*n*=33 were diagnosed with unspecified dementia).

To better isolate disease-specific effects, we investigated polygenic risk scores (PRS) indexing the genetic susceptibility to a certain disease,^41^ comparing high genetic risk (top 1%) and low genetic risk (bottom 50%) individuals (Fig. 4B). For cardiovascular disease and ischemic stroke, WMHs showed more severe alterations in high PRS individuals in anterior regions (average *β* = 0.12 and 0.18, respectively; *p* (FDR) < 0.05 for all markers except OD, T2*, QSM, and for all markers except QSM, respectively) than posterior regions (average *β* = 0.09 and 0.17, respectively; *p* (FDR) < 0.05 for WMH volume, stage, MD, FA, and for all markers except OD, respectively). For Alzheimer’s disease, WMHs showed more severe alterations in high PRS individuals in posterior regions (average *β* = 0.08; *p* (FDR) < 0.05 for stage, MD, ICVF, OD, T2*) relative to anterior regions (average *β* = 0.01; no significant associations at *p* (FDR) < 0.05). The inverse pattern of FA and OD effects in stroke and Alzheimer’s PRS echoed findings from the diagnosis analysis.

We replicated the Alzheimer’s-specific findings using ADNI3 data (Fig. 4C; demographic statistics detailed in Supplementary Table 2), which included fiber- and fluid-sensitive metrics but not myelin- and iron-sensitive metrics. Normative models were transferred to compute WMH pathophysiology maps (Methods). Compared with cognitively normal control participants (*n*=125), individuals with mild cognitive impairment due to Alzheimer’s disease (*n*=66) and individuals with Alzheimer’s disease dementia (*n*=21) showed a similar pattern of higher effect sizes in posterior WMHs, lower effect sizes for FA and higher effect sizes for OD, although some effects did not reach significance likely due to the lower sample sizes.

Finally, we quantified the replicability across the eight reported effect size patterns (Fig. 4D), observing high correlations within vascular and neurodegenerative groupings, which clustered together (Fig. 4D). This demonstrated the reproducibility of the disease-specific spatiotemporal WMH pathophysiological signatures, which clearly differed in vascular and neurodegenerative pathologies.

### Posterior WMHs connect cortical regions prone to tau pathology

Next, we investigated the grey matter correlates of our spatial WMH clusters, with the hypothesis that Wallerian degeneration stemming from grey matter Alzheimer’s pathology would affect the associated white matter fibers which could be linked to our observed pattern of fiber disorganization in posterior WMHs in neurodegenerative disorders. Using a high-resolution tractogram derived from 1001 subjects from the Human Connectome Project,^42^ we extracted streamlines passing through each white matter spatial cluster and calculated normalized cortical connectivity profiles (Fig. 5A; Methods). This showed that the posterior WMH region disproportionately connects inferior temporal and occipital regions, while the anterior WMH region connects dorsal frontal regions.

**Figure 5.**
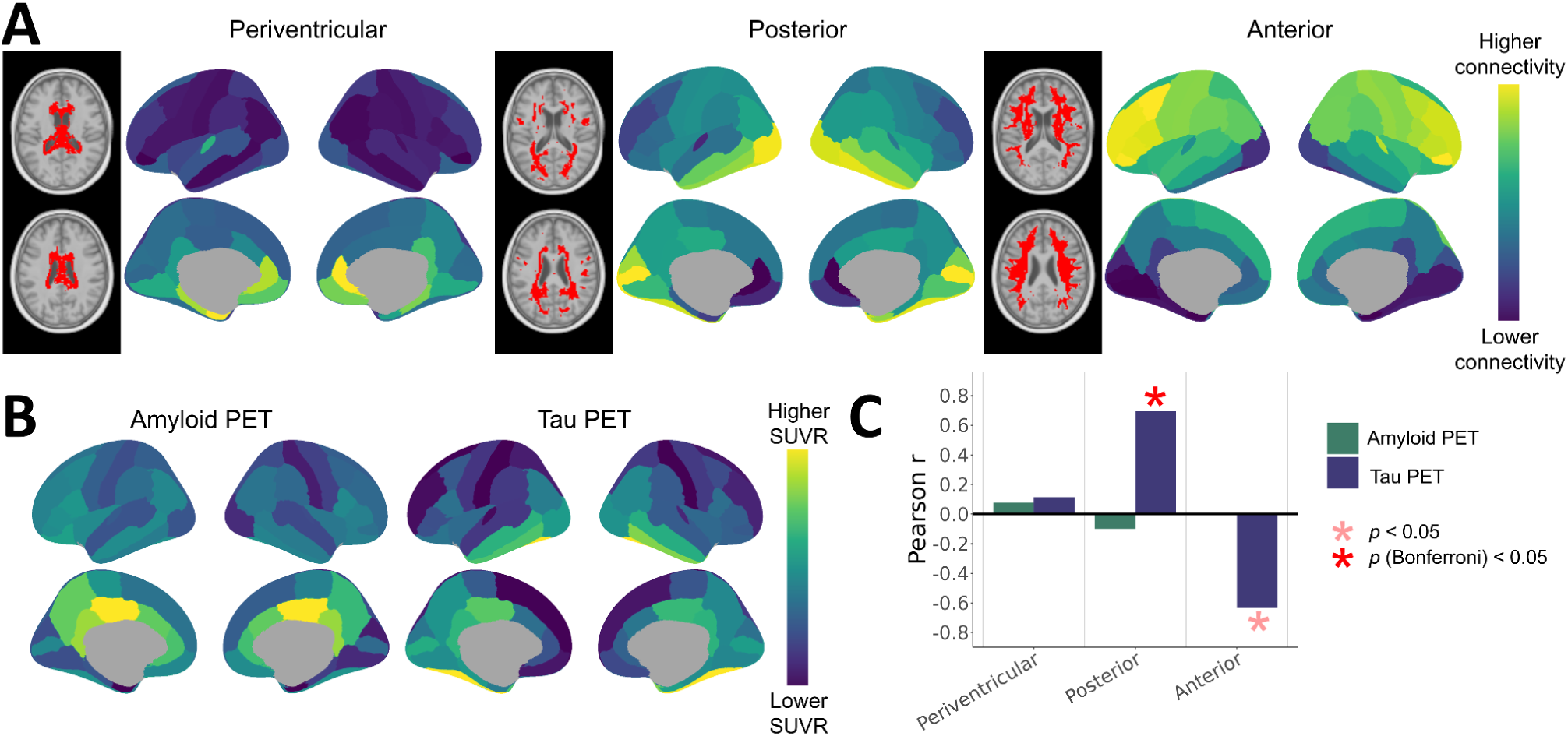
Cortical connectivity profiles of WMH regions and their associations with amyloid and tau PET distributions. **A)** The total amount of streamlines passing through the WMH spatial clusters and connecting each cortical grey matter region were calculated and normalized. Yellow regions indicate a disproportionately higher connectivity relative to the other clusters. **B)** Group averages of amyloid and tau PET SUVR distributions in amyloid-positive individuals from the PREVENT-AD cohort. **C)** Spatial Pearson correlations between cortical WMH patterns and PET averages. *p*-values were calculated from spin test null models preserving the spatial autocorrelation of cortical maps. Significant effects at the uncorrected and Bonferroni-corrected *p*<0.05 levels are indicated with asterisks in shades of red.

The cortical patterns were compared with average maps of amyloid-beta (Aβ) and tau positron emission tomography (PET) standardized uptake value ratio (SUVR) in Aβ-positive individuals from the PREVENT-AD cohort (Fig. 5B).^43^ This dataset was chosen due to the high representation of individuals at early stages in the Alzheimer’s disease spectrum. We observed high correlations between cortical connectivity profiles of the posterior WMH region and the tau PET distribution (*r*=0.65, *p* (Bonferroni) = 0.005), which was significantly different from a spatial permutation-based null (Fig. 5C).^44^ No significant associations were observed for Aβ PET. These results demonstrate that the posterior WMH region structurally connects cortical regions prone to early tau pathology, but not amyloid pathology.

## Discussion

The interpretation and clinical management of WMHs are confounded by their differing vascular or neurodegenerative pathologies. This nuance is rarely taken into account as there is no established framework or guidelines to distinguish these two processes *in vivo*. We hypothesized that the underlying pathological contributions to WMHs would be revealed by diverging spatiotemporal pathophysiological signatures captured by fluid-, fiber, and myelin- and iron-sensitive microstructural MRI metrics. Leveraging a large sample size (*n*=32,526), we estimated subject-level WMH pathophysiology and group-level spatiotemporal patterns, as well as derived disease-specific signatures in vascular and neurodegenerative disorders.

Rather than the traditional periventricular/deep WMH classification,^18^ our data-driven approach revealed that deep-anterior WMHs had more severe pathophysiology than periventricular-posterior WMHs, which does not seem to be caused by spreading patterns. This anterior-to-posterior differentiation is consistent with other data-driven evidence based on WMH co-occurrence.^12,45^ Temporally, WMHs followed a consistent pathophysiological cascade across individuals. In agreement with the prevailing hypothesis, water infiltration and inflammation seem to be early events, as reflected in the early MD abnormality.^4,25–27^ While the early reduction in ICVF could point to axonal loss,^28,29^ we speculate that inflammatory swelling of cells could increase the extracellular signal fraction and thus decrease the intracellular signal fraction, resulting in the observed effect. The combination of positive T2* abnormality, consistent with our previous work,^46^ and very low QSM positive abnormality potentially indicates myelin and iron loss, as these two processes would have an additive positive impact on T2* but would cancel each other out on QSM.^31,32^ While iron accumulation occurs in aging, especially in subcortical grey matter regions,^47^ iron loss in white matter could be caused by the death of oligodendrocytes, which contain large amounts of paramagnetic iron.^48^

We uncovered a signature of neurodegeneration-related WMHs in individuals with dementia and Alzheimer’s disease spectrum disorders, showing a posterior dominance and a reproducible pattern of specific effects on fiber-sensitive metrics. This finding adds to the body of literature linking posterior WMHs to Alzheimer’s disease.^12,13,49,50^ The pathophysiological signature points to a selective degeneration of fibers, since such process would reduce the complexity of diffusion orientations as water would diffuse relatively more strongly in the direction of the spared fibers, explaining the observed lower OD and relatively preserved FA. This interpretation is further supported by our finding that posterior WMH regions structurally connect cortical regions prone to early tau accumulation, pointing to a link between tau pathology in inferior temporal regions, degeneration of the associated fiber populations, and the appearance of posterior FLAIR hyperintensities. Cortical regions connecting through WMHs have been shown to be disproportionately affected relative to unconnected regions.^51,52^

While we cannot exclude the possibility that the neurodegenerative WMH pattern could be due to vascular dysfunction specific to Alzheimer’s disease, our interpretation is supported by both *in vivo* and *ex vivo* studies showing the absence of ischemic cerebrovascular pathology in posterior WMHs in Alzheimer’s disease^11,12,53,54^ (see reviews).^10,55^ Furthermore, we observe the same pattern in individuals at high genetic risk of Alzheimer’s disease, where vascular risk factors are likely similarly distributed relative to low genetic risk-individuals, and our findings are consistent with the reported early emergence of WMHs in autosomal-dominant Alzheimer’s disease.^49,54^ Anterior WMHs have been associated with both neurodegeneration and small vessel disease,^56^ pointing to an overlap in pathology that cannot purely be distinguished by neuroanatomical location and supporting the need for WMH pathophysiological assessments *in vivo*, since both vascular and Alzheimer’s pathology are frequently comorbid in the general population.^57^

Our framework for disentangling neurodegenerative and vascular contributions to WMHs *in vivo* has broad implications for the clinical management of patients and the interpretation of research findings and clinical trials. Hypothetically, aggressively treating hypertension following a clinical radiological finding of large WMHs may not be warranted if these are mostly related to neurodegenerative processes and increases the risk of important side effects including insomnia and vision problems.^58^ Second, the use of amyloid-clearing drugs like lecanemab is currently restricted to individuals without comorbid cerebrovascular pathology since they are at higher risk of developing potentially severe amyloid-related imaging abnormalities (ARIA).^59^ Clarifying WMH pathogenesis could improve the selection of patients for these treatments.

While our findings are robust due to the large sample size and multiple sensitivity and replication analyses, our work also has some limitations. First, the interpretation of MRI microstructural metrics is complex and may not hold in different disease contexts.^60^ In other words, there is no one-to-one mapping of any MRI metric to any specific pathophysiological process.^61^ Our multimodal approach combined metrics with different specificities and sensitivities to provide better-informed inferences of biological processes than any metric individually. In future work, we aim to confirm these biological interpretations by correlating microstructural MRI values within WMHs with histology and immunohistochemistry measures, leveraging the Douglas-Bell Canada Brain Bank which contains over 3,000 brain specimens.^62^ Second, it remains unclear if and how Alzheimer’s cortical pathology and our derived signature of neurodegeneration-related WMHs are causally linked. It will be critical to determine the direction of this association, with the possibilities of a white matter-first process (i.e., retrograde degeneration), grey matter-first process (i.e., anterograde Wallerian degeneration), or a mix of both.

## Acknowledgments

We wish to thank all UK Biobank, PREVENT-AD, and ADNI participants and staff for their invaluable contributions. This research has been conducted using the UK Biobank Resource under Application Number 45551. This research used the NeuroHub infrastructure and was undertaken thanks in part to funding from the Canada First Research Excellence Fund, awarded through the Healthy Brains, Healthy Lives initiative at McGill University. OP is funded by the Alzheimer Society of Canada and the Fonds de Recherche du Québec - Santé (FRQS). MMC receives salary support from the Fonds de Recherche Québec – Santé and from a James McGill Professorship. MMC also receives research support from Canadian Institutes of Health Research, Natural Sciences and Engineering Research Council – Canada, McGill University’s Health Brains for Health Lives (a Canada Research Excellence Fund Initiative), and TRIDENT (a New Frontiers in Research Fund program). MD receives salary support from the Fonds de Recherche Québec – Santé and research support from Canadian Institutes of Health Research, Natural Sciences and Engineering Research Council – Canada, and Brain Canada. ZA is funded by the Canada Brain Research Fund (CBRF) Rising Star program. MC is funded by the Fonds de Recherche du Québec - Santé (FRQS). AB received support from the Alzheimer Society of Canada and Healthy Brains Healthy Lives. The PREVENT-AD program was launched in 2011 as a $13.5 million, 7-year public-private partnership using funds provided by McGill University, the Fonds de Recherche du Québec – Santé (FRQ-356162), an unrestricted research grant from Pfizer Canada, the J.L. Levesque Foundation, the Douglas Hospital Research Centre and Foundation, the Government of Canada, the Canada Fund for Innovation, the Canadian Institutes of Health Research (SV: 178385, JP: 153287, 178210, LC: 165921, TS: 175328,) the Alzheimer Society of Canada, the National Institutes of Health of the United States (NS: NIH AG068563), the Alzheimer Association (SB: AARG-NTF 926696) and Brain Canada Foundation.

## Competing interests

The authors declare no competing interests

## Methods

### Study design and participants

The UK Biobank (UKB) dataset is a cohort study comprised of over 500,000 individuals from midlife to advanced age (application #45551). The study was approved by the North West Multicenter Research Ethics Committee (United Kingdom). A subset of ∼40,000 participants underwent a comprehensive brain MRI protocol. The Alzheimer’s Disease Neuroimaging Initiative (ADNI) dataset is a longitudinal observational study composed of participants along the Alzheimer’s disease spectrum (http://adni-info.org/) and was approved by the institutional review board at each participating institution. We focused on the ADNI3 dataset since it included the multi-shell diffusion MRI acquisition necessary to derive NODDI metrics. The Pre-symptomatic Evaluation of Experimental or Novel Treatments for Alzheimer Disease (PREVENT-AD) is a longitudinal study comprised of individuals who have parents or siblings diagnosed with Alzheimer’s disease and were cognitively normal at recruitment.^43^ It was approved by the institutional review board at McGill University. All participants gave written informed consent prior to participation in the studies.

### Image processing

MRI acquisition parameters for UKB and ADNI3 are available in Supplementary Methods 1. Microstructural maps were processed by the UKB team,^22,63^ and we processed ADNI microstructural maps ourselves, matching the UKB processing steps when possible. Processing steps to derive microstructural metrics from diffusion and susceptibility-weighted imaging are detailed in Supplementary Methods 2.

Since voxel-by-voxel correspondence and accurate segmentation of WMHs were crucial for our application, we updated key processing steps from UKB and applied those same steps to the ADNI dataset. T1w and FLAIR images were denoised,^64^ corrected for N3 inhomogeneities,^65^ and intensity normalized. Brain masks were calculated using the BEaST algorithm.^66^ A UKB study-specific template space was generated using the processed T1w images of 200 subjects with representative age and sex distributions (available at https://github.com/CoBrALab/WMH_patho_UKB), and an ADNI template was generated similarly from all cross-sectional T1w images.^23^ All modalities were then aligned at the subject level by performing rigid registration of other modalities (FLAIR, diffusion, and susceptibility) to T1w subject space using the Advanced Normalization Tools software.^67^ We then performed multispectral non-linear registration to the study-specific templates using preprocessed T1w images and fractional anisotropy maps supersampled to 1 mm isotropic^68^ as inputs.^67^ The ADNI template was non-linearly registered to the UKB template. All microstructural maps were transformed in UKB space using 4th-order B-spline interpolation at a 2 mm isotropic resolution to match the diffusion-weighted imaging resolution (of note, the susceptibility-weighted imaging acquisition had an original resolution of 0.8 x 0.8 x 3 mm). The rich spatial contrast of fractional anisotropy maps allowed for increased registration accuracy of white matter tracts,^69^ a crucial aspect for our application. WMHs were segmented in native space with the Brain tIssue SegmentatiON (BISON) algorithm, a multi-contrast random forest classifier^15^ that was retrained on 60 manually-labeled WMH masks on UKB data using both T1-weighted and FLAIR images as inputs.

### Quality control

The MRI data underwent multiple steps of quality control. We visually inspected T1w and FLAIR images according to guidelines established by our group and excluded scans showing significant motion artifacts (https://github.com/CoBrALab/documentation/wiki/Motion-Quality-Control-(QC)-Manual).

Motion during diffusion MRI scanning was quantified using an automated tool^70^ and scans with a mean absolute head motion (UKB field 24450) above 3 mm were excluded. Participants were further excluded if they failed any image processing steps or had a multiple sclerosis diagnosis according to the first-occurrence field (UKB field 131042).

### Normative modeling and WMH pathophysiology

Normative modeling of NAWM was performed on a voxel-wise basis in the space of our UKB template for each of the seven markers only using UKB microstructural data (Fig. 1A). In other words, only individual-level voxels that were labeled as NAWM by the BISON algorithm were included in the normative models. We used Bayesian linear regression from the PCN toolkit^34^ with the covariates of age (modeled with 3rd-order B-splines with 4 knots) and sex. Age- and sex-specific maps of average and standard deviations of NAWM microstructure were then generated and used to z-score the microstructure of individual-level voxels labeled as WMH (example in Fig. 1B). This process crucially isolates the impact of the presence of a WMH on the microstructure and removes the spatial microstructural contrast driven by normal anatomical variations.

The normative model hyperparameters were then transferred to ADNI data in order to account for site and scanner variations. The adaptation sample to estimate the site effect was comprised of the first timepoint available of cognitively normal individuals within the UKB age range (45-81 years old; *n*=114), which is deemed a large enough sample for accurate transferring of normative model hyperparameters.^71^ For the ADNI subjects older than 81 years old (*n*=35), pathophysiology maps were calculated from normative values of 81-year-olds in order to not extrapolate the models.

### Analyses

All linear models were processed in R/4.1.2 using the *lm* function, scaling all continuous variables to obtain standardized beta coefficients. *p*-values were corrected using the false discovery rate (FDR)^40^ on a per-analysis basis unless otherwise indicated.

#### Spatial clustering

Between-subject averages of WMH pathophysiology for each of the seven microstructural markers were computed (Fig. 2A). Voxels with a low prevalence of WMH (<30) or NAWM (<5000) labels were excluded (32,424 excluded voxels out of 54,257 total voxels). Included voxels were thus stable, robust estimates of the average WMH pathophysiology and still covered large portions of the white matter, permitted by the very large sample size. These maps were used as inputs to a spectral clustering algorithm using the *Spectrum* package in R^35^ resulting in spatial clusters of pathophysiologically similar WMHs (Fig. 2B). The eigengap method^35^ determined that the optimal number of clusters was 4 (Supplementary Fig. 2A); however, the three-cluster solution resulted in more spatially contiguous clusters with similar pathophysiological differentiation across clusters (Supplementary Fig. 2B). We calculated the Dice similarity coefficient (DSC) overlaps between our derived spatial cluster and other often-used parcellations (Fig. 2C): periventricular/deep (derived manually by defining a distance from the ventricles of >8 mm as deep), lobar,^72,73^ and cerebral arterial territory.^36^ We only included clustered voxels in the overlap calculations.

The clustering analysis was repeated in different subgroups: based on WMH volume (low: <=5000 mm^3^, *n*=27,511; medium: >5000 and <=10000 mm^3^, *n*=3,103; high: >10000 mm^3^, *n*=1,192), excluding voxels with prevalence of WMH<10 (Extended Fig. 1A), and across sexes, excluding voxels with prevalence of WMH<15 (Extended Fig. 2A). DSCs between the original clusters and subgroup clusters were calculated, only including voxels that were present in both clustering solutions in the calculation (Extended Fig. 1B and Extended Fig. 2B).

To derive a final parcellation covering all possible WMH locations (i.e., including voxels that were previously excluded based on low WMH or NAWM prevalence), we used a search area strategy: for each excluded voxel, we sampled the assigned clusters of the surrounding voxels and calculated the most prevalent cluster assignment. If the excluded voxel was not immediately surrounded by voxels with an assigned cluster, we extended the search area iteratively by steps of one voxel until there were voxels with an assigned cluster in the search area. This final parcellation covering the whole white matter is shown in Supplementary Fig. 3 and is made publicly available in both our custom UKB template space and in standard MNI ICBM152 09c symmetric space (https://github.com/CoBrALab/WMH_patho_UKB).

We then calculated region-of-interest measures of WMH pathophysiology by computing the median pathophysiological value for each metric within WMHs in each spatial cluster. In cases with fewer than 5 WMH voxels within a spatial cluster, data was imputed as 0, indicating no WMH-related microstructural abnormality. Region-wise WMH volumes were calculated in common space (inherently normalized for brain size) and log-transformed, as recommended.^74^ To simplify the interpretation, we inverted the values of markers that showed negative effects in WMHs (FA, ICVF, and OD) so that an increase in z-scores is consistently interpreted as worsening WMH pathophysiology for all markers.

#### Temporal modeling

To model the temporal progression of WMH pathophysiological events, we used the Subtype and Stage Inference (SuStaIn) technique,^37,75^ specifically the piecewise linear z-score model since our data was expressed as abnormality z-scores (Fig. 3A). Pathophysiological events were determined based on z-score thresholds (Z = 0.5, 1, 1.5, 2, 3, 4, 5) and the maximum z-score was determined empirically on a per-marker basis (at least 1% of participants needed to reach the threshold; Supplementary Fig. 4). SuStaIn was performed using 10-fold cross-validation with 15 startpoints and 10,000 Monte Carlo Markov Chain resamples at each fold. We compared the one-subtype solutions to the three-subtype solutions and concluded that there was no evidence of sub-trajectories (Extended Fig. 3). We also modeled the relationships between WMH pathophysiology and volume (i.e., using WMH volume as an empirical proxy of WMH temporal progression). Non-linear relationships with log-transformed WMH volume were modeled with 4th-order B-splines (Extended Fig. 4).

#### Sex differences

Sex differences in WMH measures were investigated using linear models 1) for regional WMH volumes, correcting for age (to investigate simple sex differences; Fig. 3B), and 2) for WMH pathophysiological estimates, additionally correcting for region-specific WMH volume (to investigate differences in WMH pathophysiology when males and females have equivalent extents of WMHs; Fig. 3C).

#### Associations with vascular and neurodegenerative disorders

UK Biobank diagnosis data was used through the first occurrences field (category #1712) which mapped data from self-reported medical conditions, primary care, hospital inpatient, and death register records to ICD-10 codes together with a date of diagnosis. Control subjects were defined as subjects with no endocrine, circulatory, behavioral, nervous, or metabolic disease-related lifetime diagnoses (*n*=10,629), with some exceptions for high-prevalence diagnoses that we did not expect to have a significant impact on WMHs: depressive episode (F32; *n*=4,163), migraine (G43, *n*=2,632), other anxiety disorders (*n*=2,566), hemorrhoids (I84, *n*=1859). Case subjects were defined by groupings of ICD-10 codes (detailed in Supplementary Fig. 6) regardless of the timing relative to the MRI visit. We then compared WMHs between cases and controls using linear models while covarying for age and sex (Fig. 4A).

We investigated the associations between WMH pathophysiology and genetic susceptibility to vascular and neurodegenerative diseases, as indexed with polygenic risk scores (PRS) computed by UK Biobank.^41,76^ We used the standard PRS calculated on all UK Biobank participants. Briefly, the authors used genome-wide association study (GWAS) meta-analysis results external to UK Biobank to train trait-specific PRS algorithms and then calculated subject-wise PRS values as the sum of the per-variant effect size multiplied by allele dosage. The first 5 genetic principal components were used to obtain distributions with approximately zero means and unit variances per ancestry group. We used a similar case-control design as the diagnosis analysis, where controls were defined as individuals in the bottom 50% of a PRS and cases as individuals above the top 1% of a PRS. Groups were compared using linear models with no covariates (Fig. 4B).

For the ADNI replication analysis, our sample was composed of cognitively normal participants (*n*=125), participants with a diagnosis of mild cognitive impairment determined to be caused by Alzheimer’s disease (*n*=66), and participants with a diagnosis of Alzheimer’s disease dementia (*n*=21). We kept the last timepoint available with the required MRI acquisitions for each participant in order to maximize the number of individuals diagnosed with mild cognitive impairment or dementia. Groups were compared with linear models controlling for age and sex (Fig. 4C). Amyloid and tau PET data availability in ADNI was insufficient to investigate associations with WMH markers with adequate statistical power (Amyloid PET: 56 controls, 19 MCI, 10 AD; tau PET: 43 controls, 20 MCI, 9 AD).

Each effect size pattern was correlated to all other patterns with Pearson’s correlations, excluding modalities that were not present for the correlations with ADNI patterns. Hierarchical clustering was performed on the resulting correlation matrix using the *hclust* function in R (Fig. 4D).

#### Cortical grey matter connectivity profiles

To investigate the cortical regions that were structurally connecting through our three WMH spatial clusters, we used a high-resolution tractogram of 10 million streamlines derived from the Human Connectome Project dataset with diffusion MRI-based tractography.^42,77^ We extracted the streamlines passing through each cluster using mrtrix3’s *tckedit* command.^78^ Using the CerebrA atlas (a precisely-registered and manually-corrected version of the MindBoggle101 atlas in MNI ICBM152 space),^79^ connectivity matrices were constructed and summed (to calculate the total connectivity of each ROI) for each of the three streamline sets using the *tck2connectome* command. Since larger ROIs will have more streamlines, and thus higher raw connectivity metrics, we further normalized the values by dividing by the number of streamlines of each ROI across all three clusters (Fig. 5A).

To calculate Alzheimer’s disease pathology maps, we used the PREVENT-AD dataset, a longitudinal cohort of individuals with a parental history of Alzheimer’s disease who were cognitively unimpaired at recruitment,^43^ and calculated average maps of amyloid-beta (Aβ) and tau positron emission tomography (PET) standardized uptake value ratio (SUVR) in Aβ positive individuals using cross-sectional data from the last available timepoint (*n*=35). Aβ PET imaging was performed with the [^18^F]NAV4694 (NAV) tracer and tau PET was performed with the [^18^F]AV1451 (flortaucipir) tracer. Processing details are available in McSweeney et al., 2020.^80^ Briefly, PET images were registered to each subject’s T1w scan, masked to exclude CSF signal, and smoothed with a 6 mm^3^ Gaussian kernel. Standardized uptake value ratios (SUVRs) were normalized with cerebellum grey matter values for Aβ PET^81^ and inferior cerebellum grey matter values for tau PET.^82^ Aβ positivity was determined based on global Aβ SUVR in a data-driven fashion using Gaussian mixture modeling with two components.^81^ PET volumetric maps were registered to MNI space and averaged. ROI values were extracted using the CerebrA atlas (Fig. 5B).^79^

We computed Pearson correlations between the cortical connectivity maps and averaged PET maps, performed “spin tests” to calculate 10,000 permutation-based nulls preserving the spatial autocorrelation,^44^ and derived *p*-values using the *neuromaps* package in Python (Fig. 5C).^83^ *p*-values were corrected with Bonferroni instead of FDR due to the lower amount of comparisons (*n*=6).

## Data availability

Data from UK Biobank is publicly available via material transfer agreements (https://www.ukbiobank.ac.uk/enable-your-research/register). Data from ADNI is publicly available via a data-sharing application (https://adni.loni.usc.edu/data-samples/adni-data/#AccessData). Data from PREVENT-AD is publicly available via a data user agreement (https://openpreventad.loris.ca/). We share all analysis code, results and figures in raw form, and our derived UKB template space and WMH parcellation based on pathophysiology on GitHub (https://github.com/CoBrALab/WMH_patho_UKB).

## Extended data

**Extended Figure 1.**
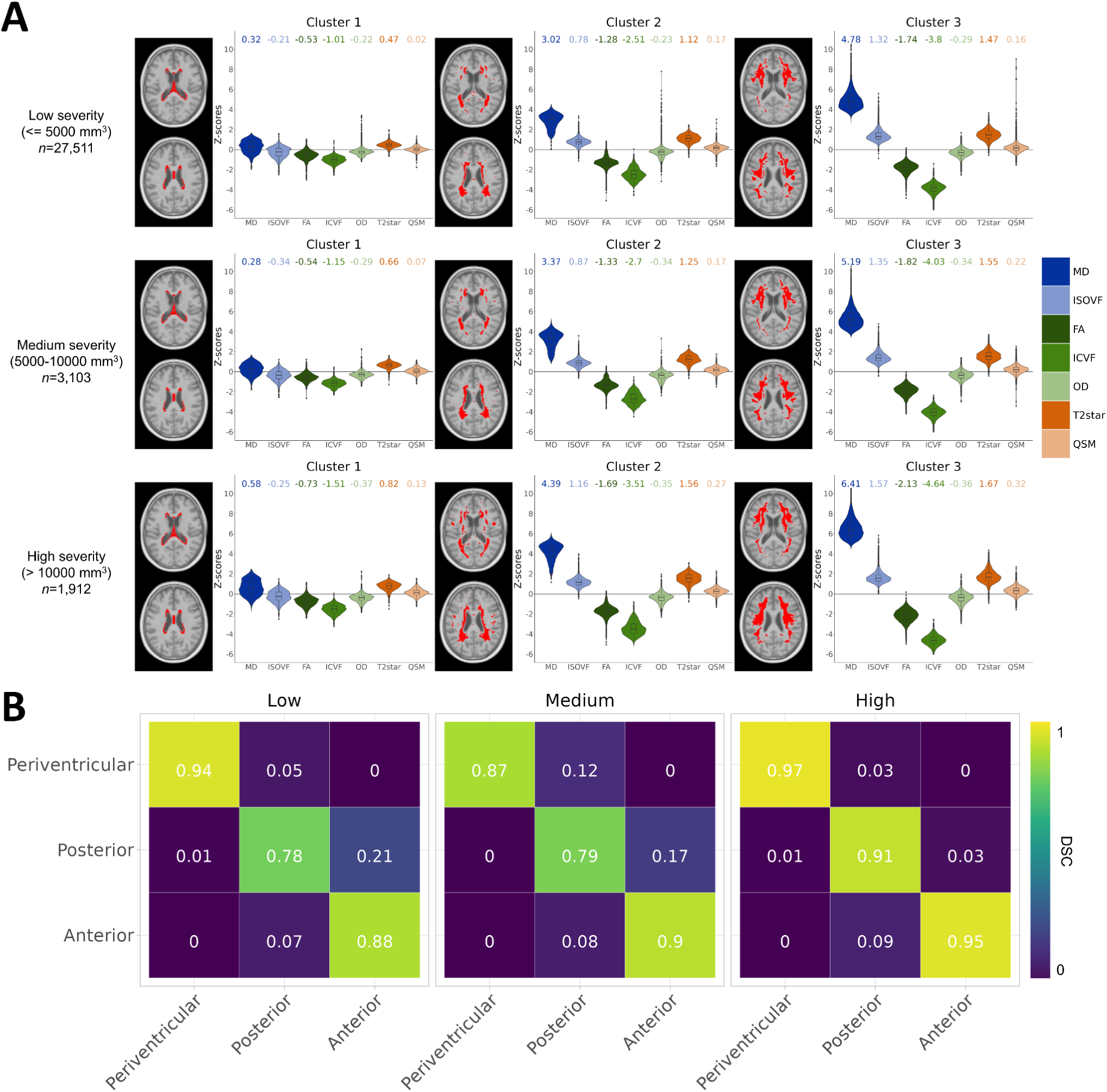
Spatial clusters by severity of WMH burden. **A)** Spatial clusters were calculated in different brackets of total WMH volumes. The sample size in each bracket is indicated. Only voxels with WMH prevalence >10 are included per group. Left: voxels included in the cluster are indicated in red. Right: Pathophysiological distributions of the voxels included in the cluster. Median values of those distributions are shown at the top. **B)** DSCs between the clusters in each severity bracket (x-axis) and the original clusters derived in the whole sample (y-axis). Only voxels included in the original and new clustering solutions are included in the calculation.

**Extended Figure 2.**
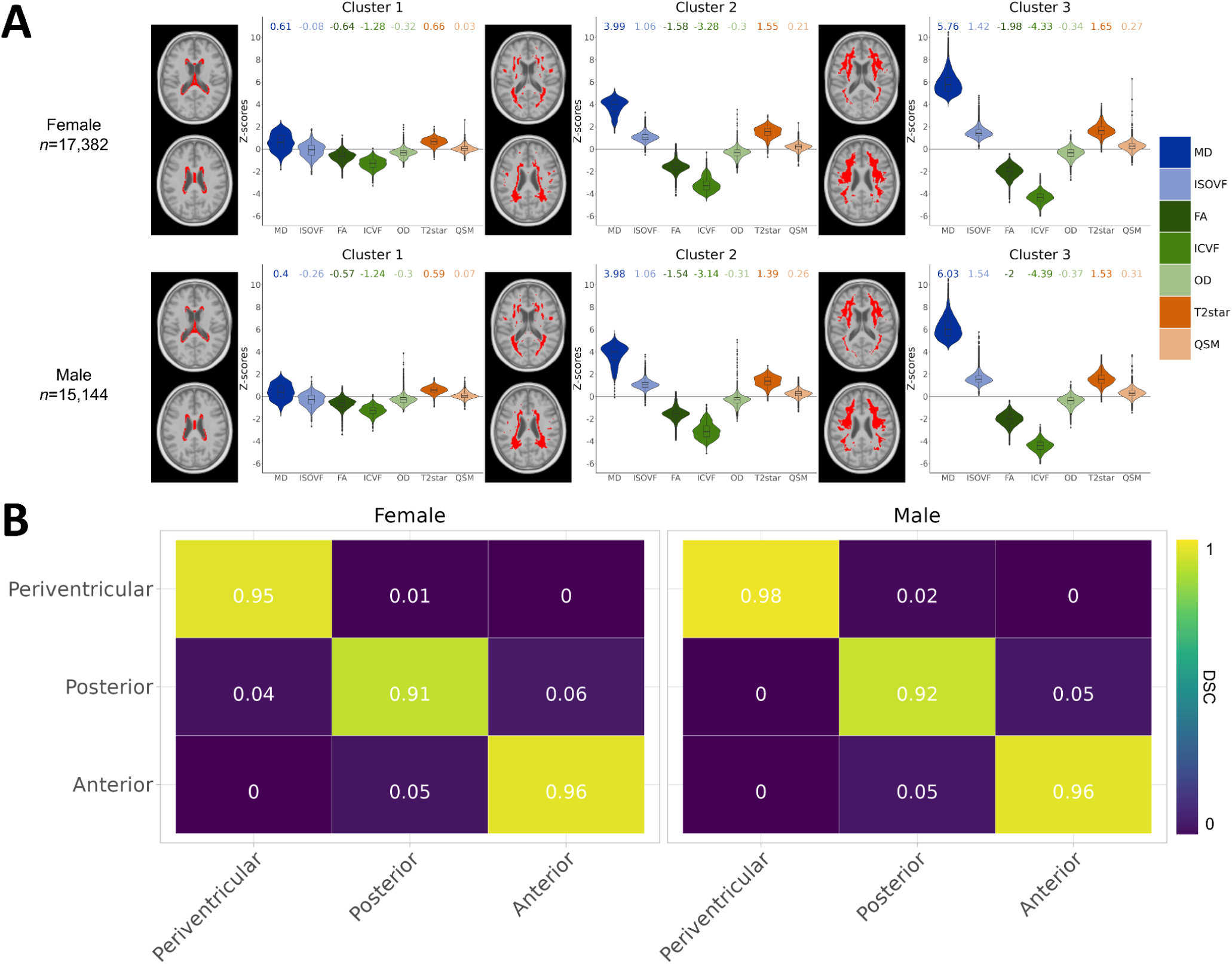
Spatial clusters by sex. **A)** Spatial clusters were calculated in females and males separately. The sample size for each sex is indicated. Only voxels with WMH prevalence >15 are included per group. Left: voxels included in the cluster are indicated in red. Right: Pathophysiological distributions of the voxels included in the cluster. Median values of those distributions are shown at the top. **B)** DSCs between the clusters calculated in females and males (x-axis) and the original clusters derived in the whole sample (y-axis). Only voxels included in the original and new clustering solutions are included in the calculation.

**Extended Figure 3.**
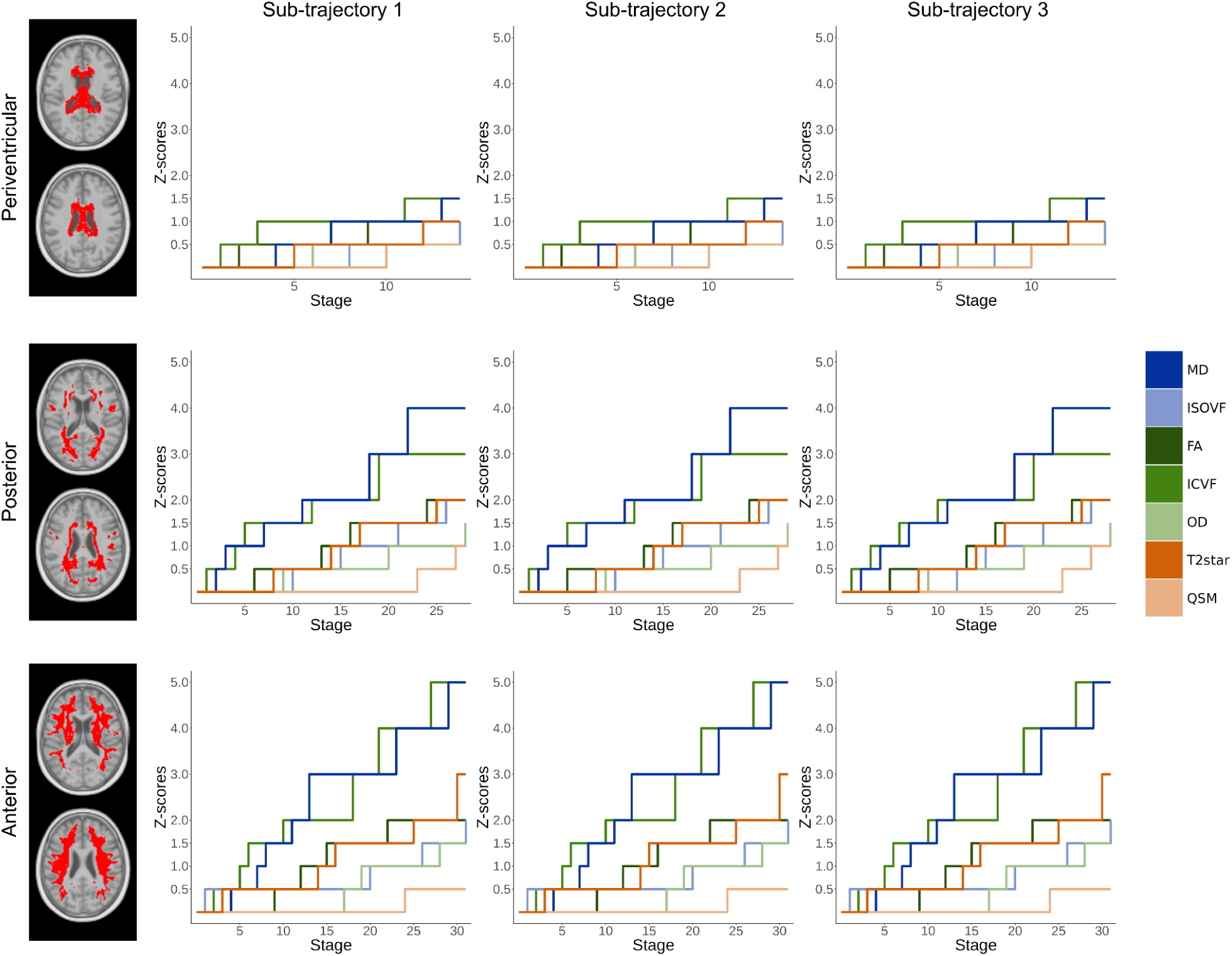
Absence of sub-trajectories of pathophysiological cascades. Within each spatial region (rows), SuStaIn was set to model three sub-trajectories (columns). Shown here are the winner-take-all trajectories. The x-axes represent a data-driven temporal axis of pathophysiological events (stages) and the y-axes represent the abnormality z-score thresholds. These sub-trajectories do not show meaningful differences when compared within regions.

**Extended Figure 4.**
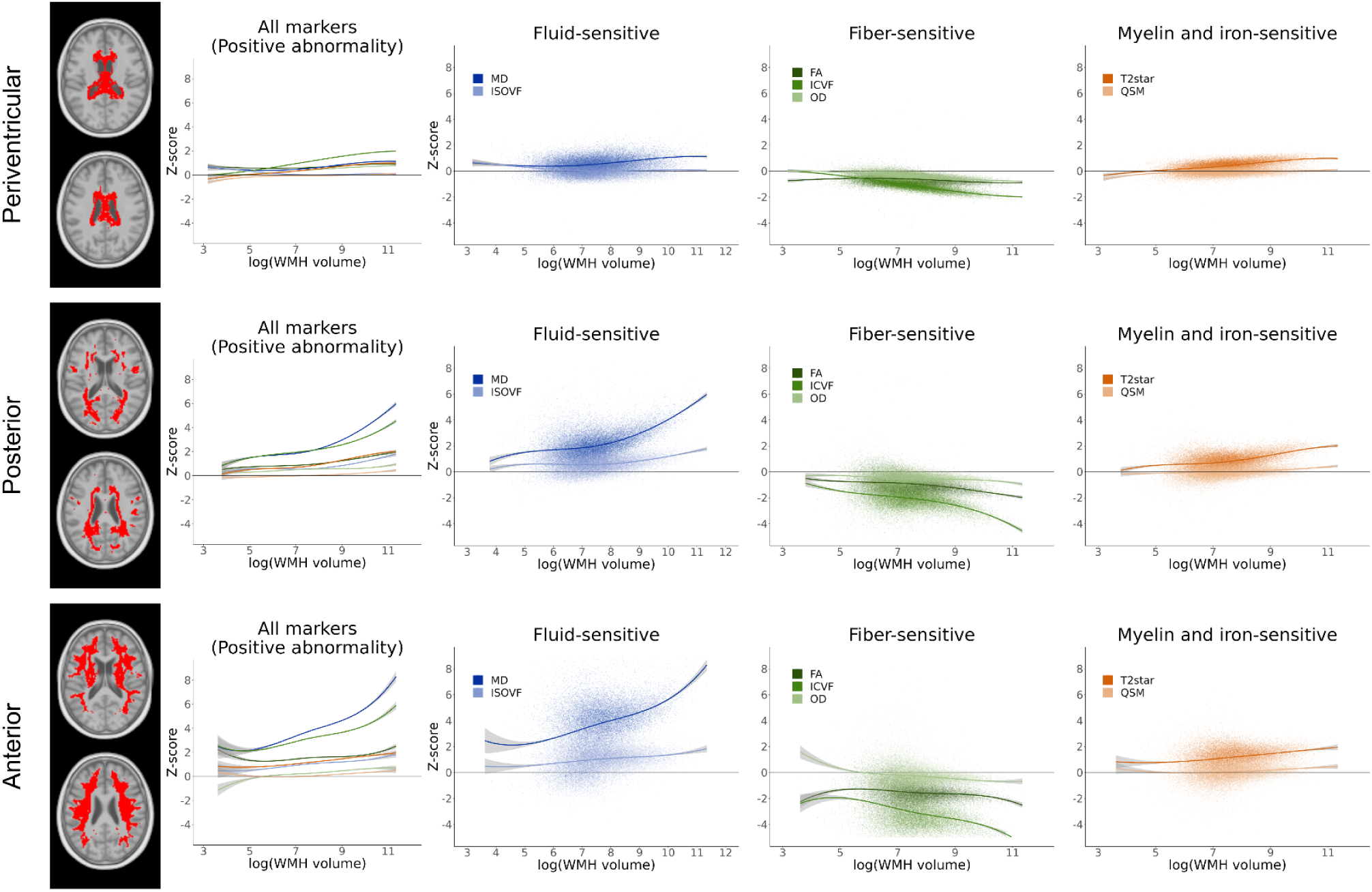
Relationships between pathophysiological estimates and WMH volume. The first graphs on the left show the trajectories of pathophysiological estimates relative to WMH volume (log-transformed) fit with fourth-order B-splines, with a higher positive abnormality indicative of worst WMH pathophysiology to be comparable with the SuStaIn results. The three graphs on the right show the trajectories by biological sensitivity, together with the distributions of the underlying data and keeping the original directionality of the measures.

## Notes

### Competing Interest Statement

The authors have declared no competing interest.

### Author Declarations

The institutional review board of McGill University gave ethical approval for this work

