## Supplementary for "Characterizing spatiotemporal white matter hyperintensity pathophysiology in vivo to disentangle vascular and neurodegenerative contributions"

### Supplementary Material

### Supplementary Methods

#### Supplementary Methods 1. MRI acquisition parameters

MRI acquisition parameters for UK Biobank

1. **T1-weighted**: Sagittal 3D MPRAGE; in-plane acceleration factor (R) = 2; inversion time (TI) = 880 ms; repetition time (TR) = 2000 ms; resolution = 1 x 1 x 1 mm
2. **T2-weighted Fluid-attenuated inversion recovery (FLAIR)**: Sagittal 3D SPACE; R = 2; partial Fourier (PF) = 7/8; fat saturation; TI = 1800 ms; TR = 5000 ms; elliptical k-space scanning; resolution = 1.05 x 1 x 1 mm
3. **Diffusion-weighted imaging (DWI)**: SE-EPI; multiband factor (MB) = 3; R = 1; TE = 92 ms; TR = 3600 ms; PF = 6/8; fat saturation; b-values: 5 x b = 0 s/mm2, 50 x b = 1000 s/mm2, 50 x b = 2000 s/mm2 (100 distinct diffusion directions); phase-encoding reversed data acquired; resolution = 2 x 2 x 2 mm
4. **Susceptibility-weighted imaging (SWI)**: Axial 3D GRE; R = 2, PF = 7/8; TE1 = 9.4 ms; TE2 = 20 ms; TR = 27 ms; resolution = 0.8 x 0.8 x 3 mm

MRI acquisition parameters for ADNI

1. **T1-weighted**: MPRAGE; acceleration factor = 2; TE = min full echo; TR = 2300 ms; TI = 900 ms; resolution = 1 x 1 x 1 mm
2. **T2-weighted FLAIR:** 3D FLAIR; TE = 119 ms; TR = 4800 ms; TI = 1650 ms; resolution = 1.2 x 1 x 1 mm
3. **DWI**: TE = 71 ms; TR = 3300 ms; b = 500, 1000, 2000 s/mm2 (112 distinct diffusion directions); resolution = 2 x 2 x 2 mm

#### Supplementary Methods 2. Processing of microstructural markers

Microstructural markers were processed by the UK Biobank team.[1,2] The multi-shell diffusion-weighted images were corrected for susceptibility artifacts using the *topup* FSL command,[3] eddy currents, head motion, and outliers using the *eddy* command,[4] and gradient distortions.[5] The cleaned first diffusion shell was used as input to the *DTIFIT* tool to generate diffusion tensor imaging (DTI) markers.[6] The multi-shell acquisition allowed for more advanced modeling techniques of the diffusion signal. Neurite orientation dispersion and density imaging (NODDI) markers were generated with the *AMICO* tool.[7,8]

The susceptibility-weighted images were saved as magnitude and phase images separately for each coil. Magnitude data was combined across coils using a sum-of-squares calculation, and from this data, T2* is calculated as the inverse of the log ratio of the two echo times scaled by the echo time difference.[1] Phase data was combined across coils using *MCPC-3D-S*, which removes phase cancellation artifacts from each echo.[9] This data underwent phase unwrapping using a Laplacian algorithm,[10,11] background field removal using *V-SHARP*,[12] and brain mask erosion to exclude voxels with low phase reliability. Dipole inversion using *iLSQR* was then used to estimate quantitative susceptibility mapping (QSM) maps,[13] which were further normalized by the subject-wise median value in the ventricles. The complete detailed processing for the DTI, NODDI, and T2* markers is available in Alfaro-Almagro et al., 2018,[1] and for QSM in Wang et al., 2022.[2]

In ADNI, we performed image processing for microstructural maps ourselves, matching the processing steps of UKB whenever possible. Since there was no phase-encoding reversed data acquired for the ADNI DWI acquisition, we corrected diffusion distortion artifacts using the Synthesized b0 for diffusion distortion correction (*Synb0-DisCo*) tool, which used deep learning to generate an undistorted b0 image using information from the T1w image.[14] This data was then fed into *topup*, *eddy*, *DTIFIT*, and *AMICO* while matching input parameters from the UKB processing.

### Supplementary Tables

| | | Complete sample (*n*=39,676) | After exclusions (*n*=32,526) |
|---|---|---|---|
| **Age** | | | |
| | Mean (s.d.; min-max) | 63.62 (7.55; 44 - 82) | 63.52 (7.49; 45 - 81) |
| **Sex** | | | |
| | Female/Male | 20,990 (52.9%)/ 18,676 (47.1%) | 17,382 (53.4%)/ 15,144 (46.6%) |
| **Ethnicity** | | | |
| | White | 38,392 (96.8%) | 31,533 (96.9%) |
| | Black | 254 (0.6%) | 197 (0.6%) |
| | Asian | 535 (1.3%) | 408 (1.3%) |
| | Mixed | 178 (0.4%) | 147 (0.5%) |
| | Other | 201 (0.5%) | 155 (0.5%) |
| **Education** | | | |
| | 7 years | 2,481 (6.3%) | 1,979 (6.1%) |
| | 10 years | 5,144 (13%) | 4,229 (13%) |
| | 13 years | 2,383 (6%) | 1,938 (6%) |
| | 15 years | 4,411 (11.1%) | 3,645 (11.2%) |
| | 19 years | 5,804 (14.6%) | 4,757 (14.6%) |
| | 20 years | 19,304 (48.7%) | 15,868 (48.8%) |

**Supplementary Table 1. UK Biobank descriptive statistics.**
Demographic statistics in the UK Biobank before and after exclusions.

| | | Complete sample (*n*=212) | Cognitively normal (*n*=125) | Mild cognitive impairment (*n*=66) | Alzheimer's disease (*n*=21) |
|---|---|---|---|---|---|
| **Age** | | | | | |
| | Mean (s.d.; min-max) | 73.41 (8.44; 51 - 93) | 72.49 (8.64; 51 - 92) | 73.86 (7.99; 55 - 93) | 77.42 (7.62; 60 - 89) |
| **Sex** | | | | | |
| | Female/Male | 123 (58%)/89 (42%) | 83 (66.4%)/42 (33.6%) | 30 (45.5%)/36 (54.5%) | 10 (47.6%)/11 (52.4%) |
| **Ethnicity*** | | | | | |
| | White | 167 (78.8%) | 96 (76.8%) | 55 (83.3%) | 16 (76.2%) |
| | Black | 29 (13.7%) | 18 (14.4%) | 7 (10.6%) | 4 (19%) |
| | Asian | 8 (3.8%) | 6 (4.8%) | 2 (3%) | 0 (0%) |
| | Mixed | 3 (1.4%) | 1 (0.8%) | 1 (1.5%) | 1 (4.8%) |
| **Education years** | | | | | |
| | Mean (s.d.; min-max) | 16.13 (2.43; 11 - 20) | 16.52 (2.35; 11 - 20) | 15.74 (2.45; 12 - 20) | 15 (2.35; 12 - 18) |
| **Amyloid status*** | | | | | |
| | Positive | 70 (42.2%) | 28 (28%) | 26 (53.1%) | 16 (94.1%) |

#### Supplementary Table 2. ADNI descriptive statistics.

Demographic statistics in ADNI in the complete sample and by cognitive group. *Removing missing values

### Supplementary Figures

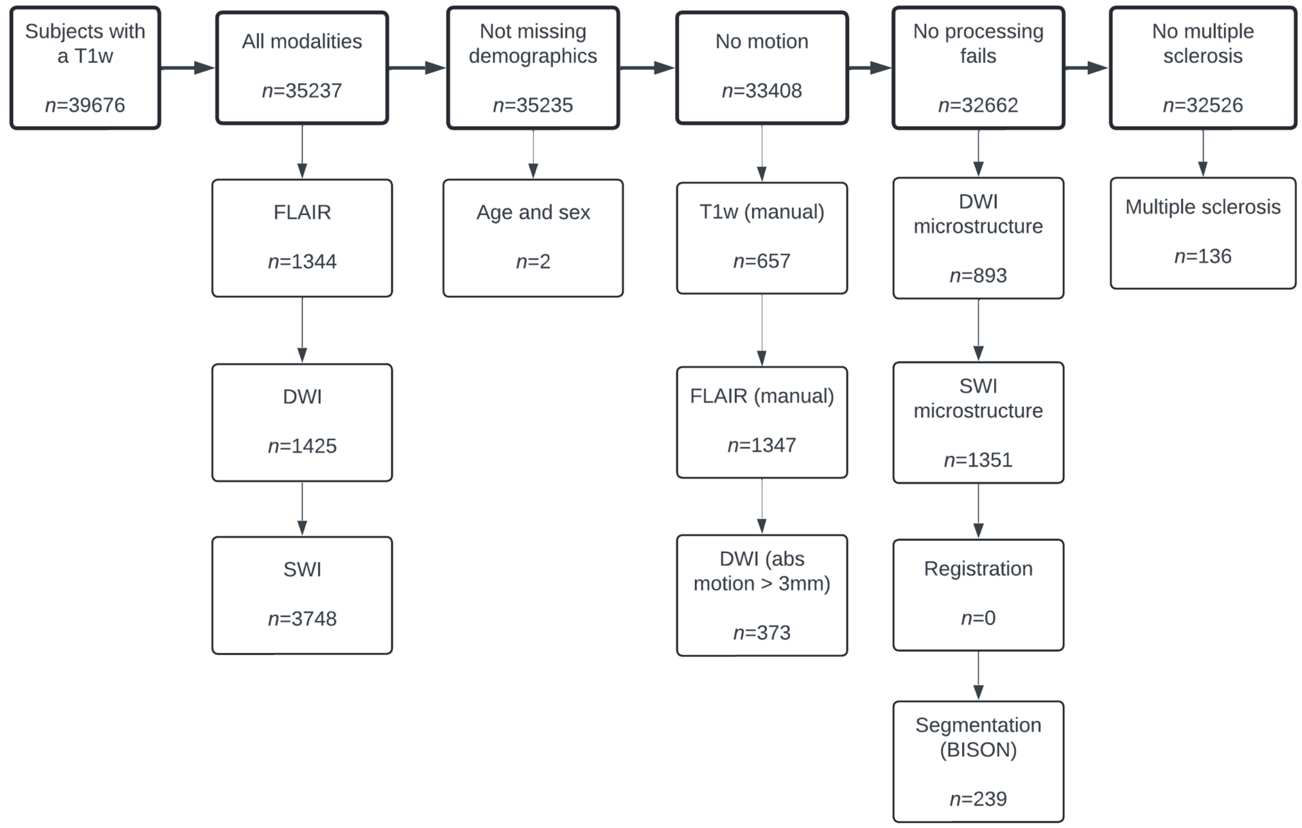

#### Supplementary Figure 1. Step-by-step exclusions.

Top rows: number of participants left after each exclusion step. Borrow rows: number of participants excluded for each criterion. Abbreviations: diffusion-weighted imaging (DWI), susceptibility-weighted imaging (SWI), T1-weighted (T1w), fluid-attenuated inversion recovery (FLAIR), BraIn SegmentatiON algorithm (BISON)

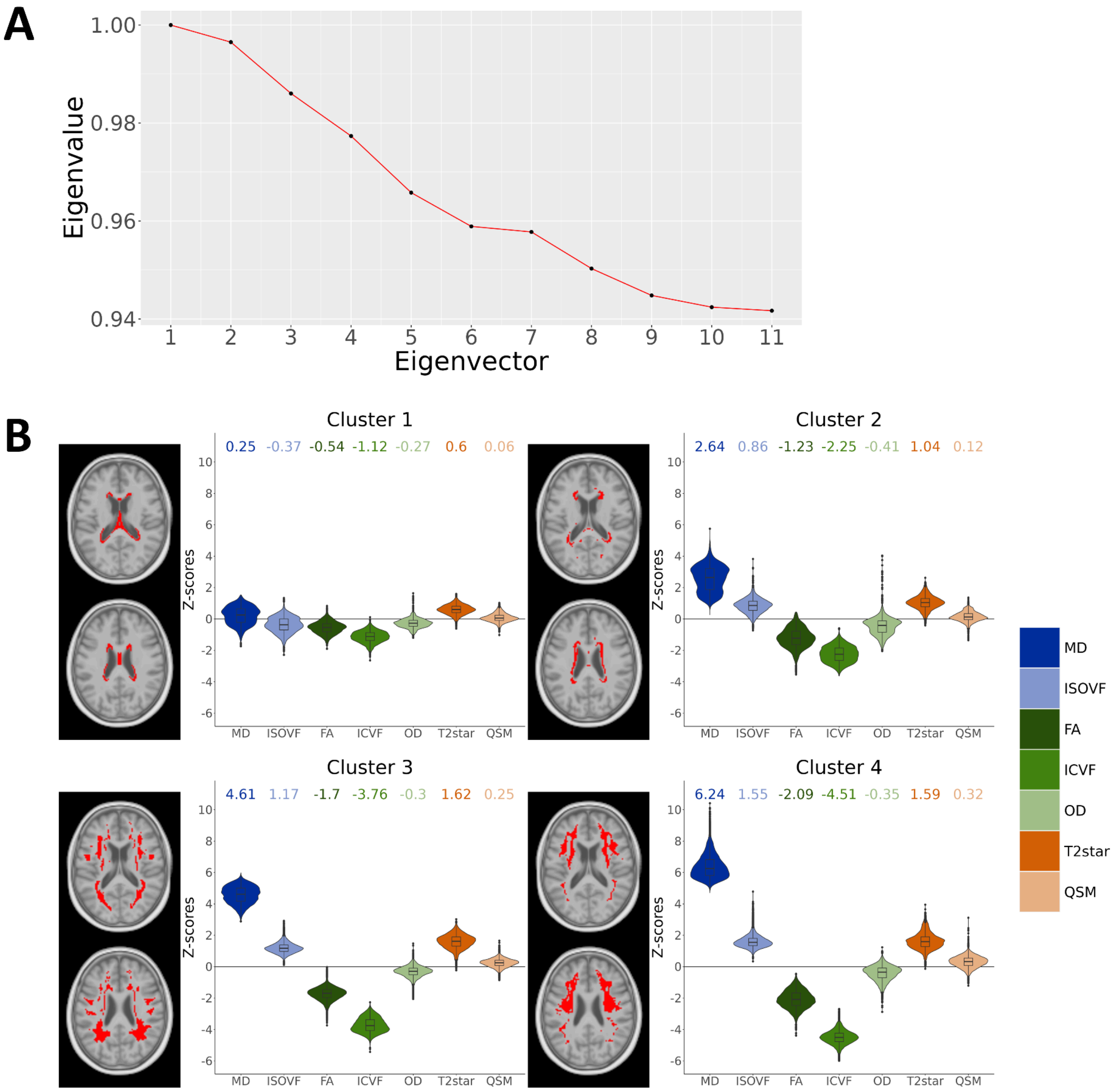

**Supplementary Figure 2. Choice of the number of spatial components and four-cluster solution.**

**A)** Eigengap graph to guide the choice of the number of clusters. According to this heuristic, the optimal number of clusters is 4. **B)** Spatial clusters for the four-cluster solution. Left: Voxels included in the cluster are indicated in red. Right: Pathophysiological distributions of the voxels included in the cluster. Median values of those distributions are shown at the top. These clusters are less spatially contiguous than the three-cluster solution, as clusters 1 and 2 are very similar.

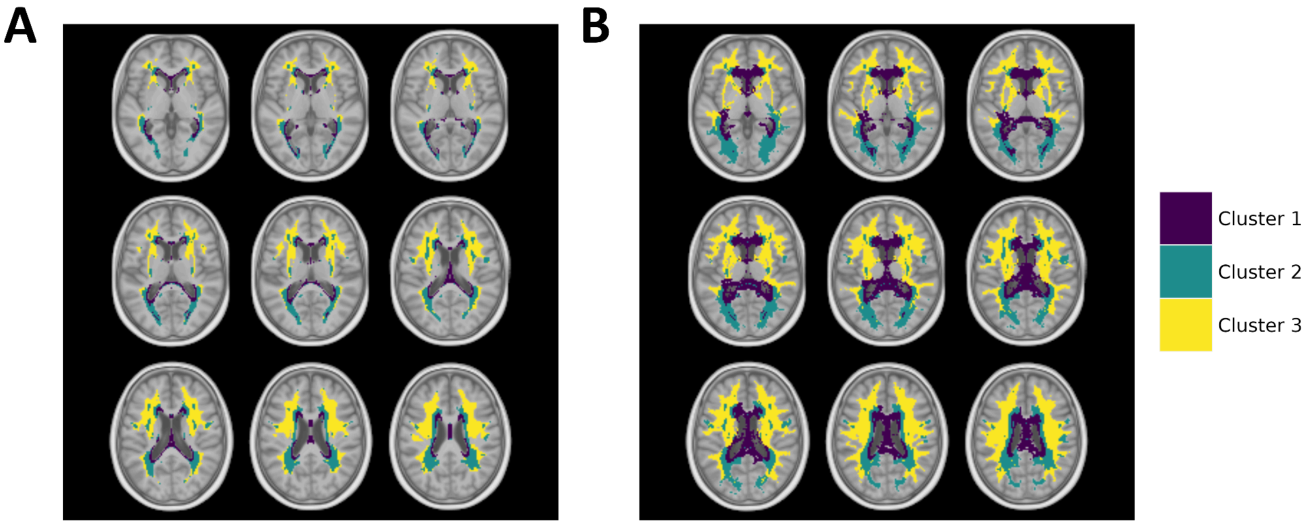

**Supplementary Figure 3. Final WMH parcellation.**

**A)** Three-cluster solution only including voxels with a high prevalence of WMHs (>30) and NAWM (>5000). **B)** Final parcellation after including low-prevalence voxels using a search area strategy

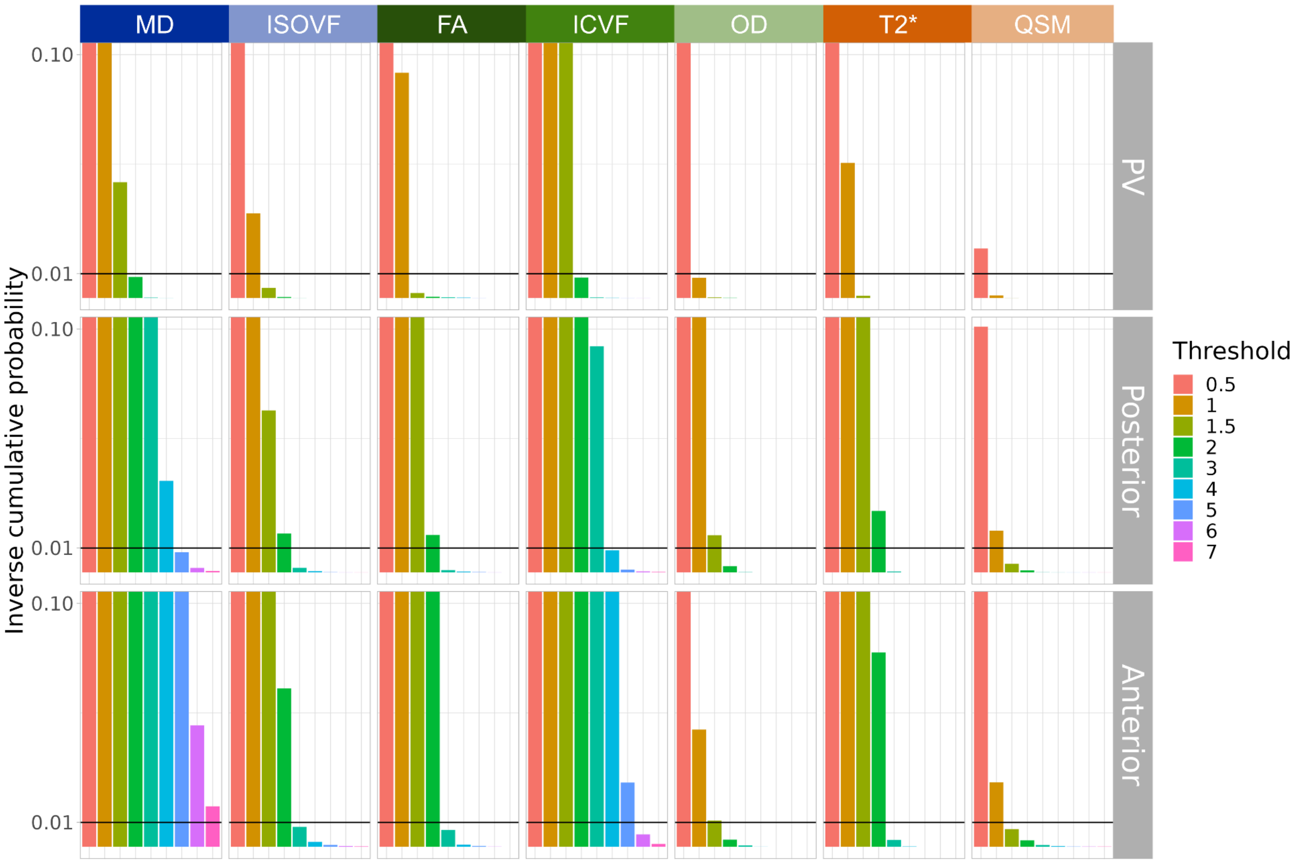

**Supplementary Figure 4. Determining z-score thresholds for SuStaIn.**

The inverse cumulative probabilities for every microstructural marker in every WMH region are shown. The maximum SuStaIn input events are determined as the last thresholds reached by at least 1% of participants (last column above the black line).

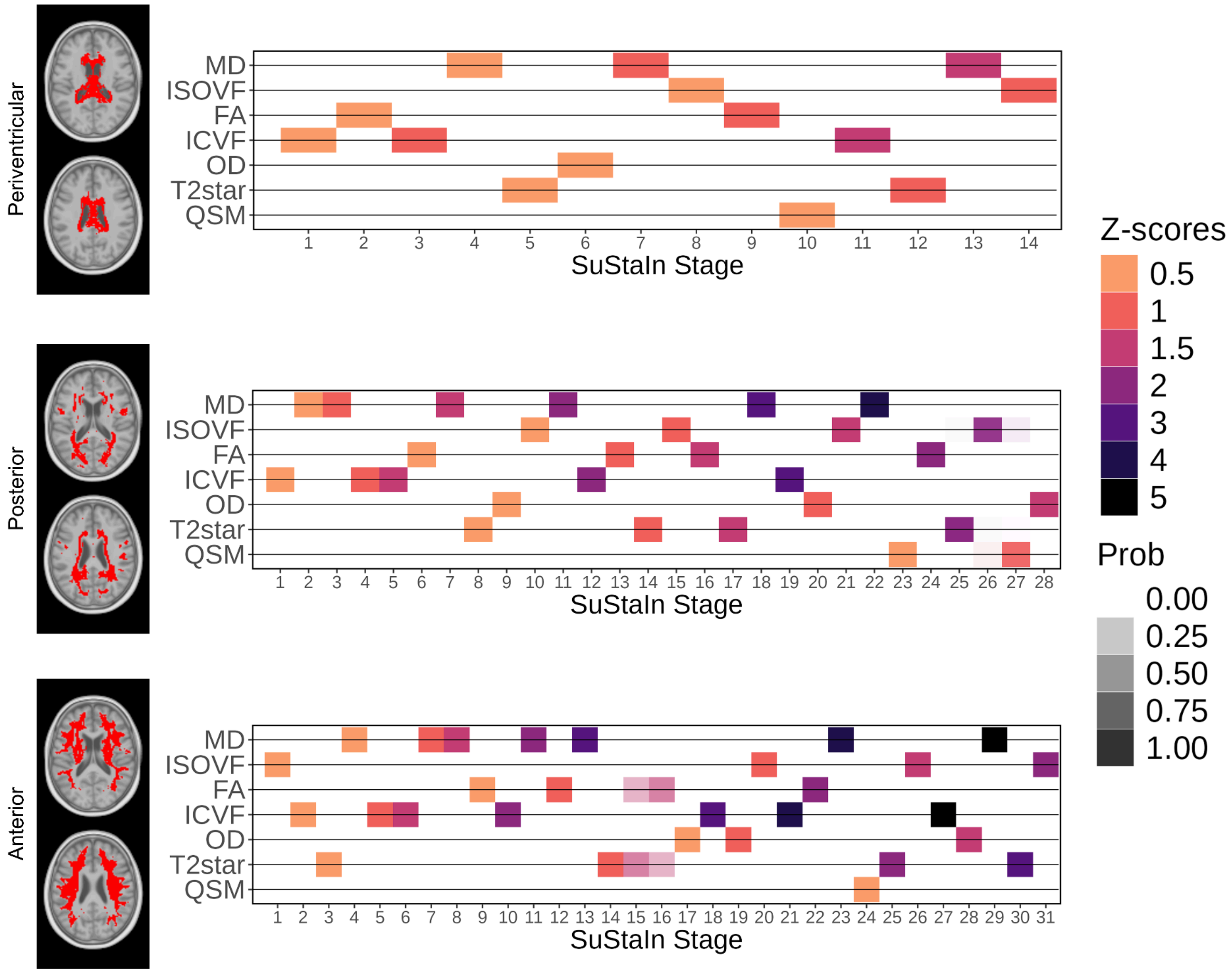

**Supplementary Figure 5. Visualizing the uncertainty in SuStaIn temporal sequences.**

Positional variance diagrams for SuStaIn trajectories. The opacity indicates the percentage of times that the pathophysiological event was placed at that stage across 10-fold cross-validation and 10,000 Monte Carlo Markov Chain resamples at each fold.

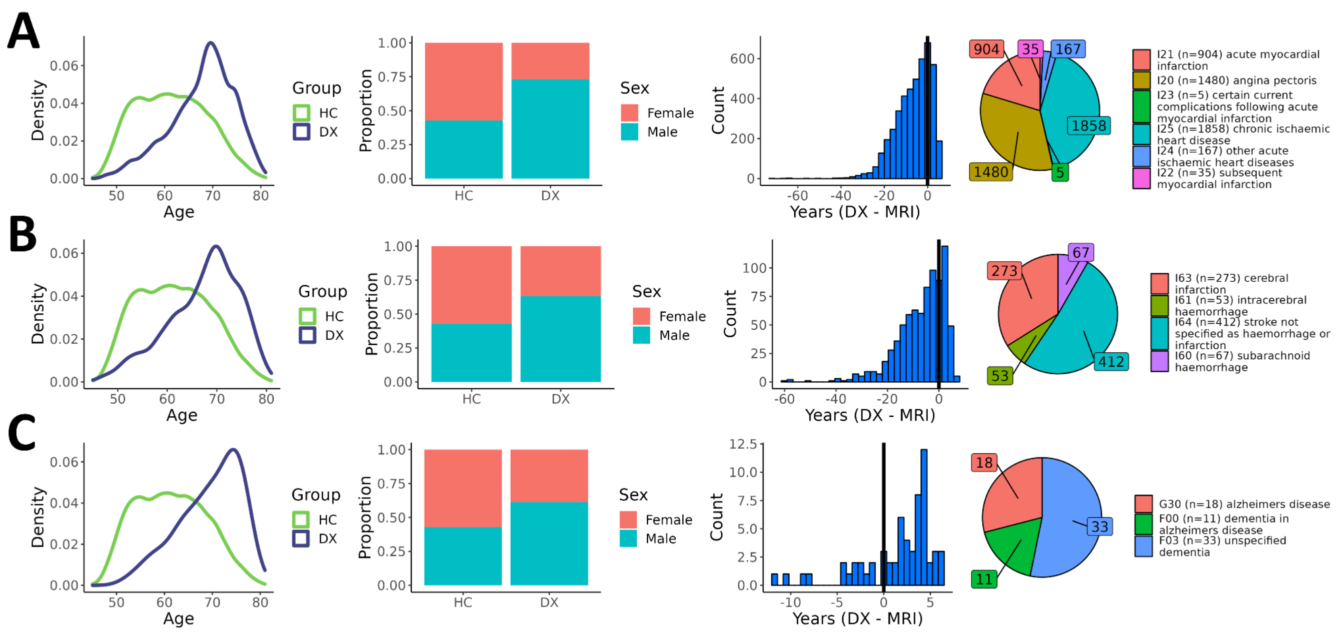

**Supplementary Figure 6. Descriptive distributions of diagnostic groupings.**

Distributions of age and sex between cases and controls, timing of the diagnosis relative to the MRI visit date, and individual ICD-10 diagnoses. **A)** Ischemic heart diseases (*n*=2,414). **B)** Stroke (*n*=645). **C)** Dementia (excluding vascular dementia; *n*=47).
